# Assessment of Bias in Clinical Trials with LLMs Using ROBUST-RCT: A Feasibility Study

**DOI:** 10.1101/2025.08.12.25333520

**Authors:** Pedro Rodrigues Vidor, Yohan Casiraghi, Adolfo Moraes de Souza, Maria Inês Schmidt

## Abstract

**BACKGROUND:** Bias assessment is a crucial step in evaluating evidence from randomized controlled trials. The widely adopted Cochrane RoB 2, designed to identify these issues, is complex, resource-intensive, and unreliable. Advances in artificial intelligence (AI), particularly in the field of large language models (LLMs), now allow the automation of complex tasks. While prior investigations have focused on whether LLMs could perform assessments with RoB 2, integrating technologies does not resolve the intrinsic methodological issues of the instrument. This is the first feasibility study to evaluate the reliability of ROBUST-RCT, a novel bias assessment tool, as applied by humans and LLMs.

**METHODS:** A sample of RCTs of drug interventions was screened for eligibility. Reviewers working independently used ROBUST-RCT to assess different aspects of the studies and then reached a consensus through discussion. A chain-of-thought prompt instructed four LLMs on how to apply ROBUST-RCT. The primary analysis used Gwet’s AC2 coefficient and benchmarking to assess inter-rater reliability of the “judgment set”, defined as the series of final assessments for the six core items in the ROBUST-RCT tool.

**RESULTS:** 54 assessments of each LLM were compared to human consensus in the primary analysis. Gwet’s AC2 inter-rater reliability ranged from 0.46 to 0.69. With 95% confidence, three of the four tested LLMs achieved ’moderate’ or higher reliability based on probabilistic benchmarking. A secondary analysis also found a Fleiss’ Kappa of 0.49 (95% CI: 0.30 – 0.60) between human reviewers before consensus, numerically higher than the values reported in prior literature about RoB 2.

**CONCLUSION:** Large Language Models (LLMs) can effectively perform risk-of-bias assessments using the ROBUST-RCT tool, enabling their integration into future systematic review workflows aiming for enhanced objectivity and efficiency.

## INTRODUCTION

Although considered the gold standard of clinical research, randomized controlled trials (RCTs) are not immune to bias. Currently, biases in RCTs are usually assessed using Cochrane’s Risk of Bias 2 (RoB 2)[1], which has well-documented limitations: its application is time-consuming[2] , complex even for experienced researchers[3], and results in low inter-rater reliability[2,4].

The automation of such tasks has been proposed for years[5]. It’s now closer to reality with artificial intelligence (AI) — particularly Large Language Models (LLMs) — in the health sciences[6–10]. At the time of this writing, the registration form for the systematic review protocol database PROSPERO[11] already includes the word “machine” in all answers to report the number of people that will assess risk of bias[12]. However, most of the literature on the use of LLMs for risk-of-bias assessment has focused on RoB 2, already known for its well-documented limitations.

The introduction of a new instrument can help to overcome these fundamental weaknesses. Designed by a panel of epidemiologists and methodologists, ROBUST-RCT[13] aims to integrate ease of use and rigor. The tool’s core items were selected based on criteria such as whether there is evidence for its influence in the effect estimation and the frequency with which the corresponding bias affects RCTs. Its development included a usability test with junior reviewers[13], emphasizing the goal of being simpler than previous tools.

Therefore, a critical opportunity lies in combining technological advancements with newer instruments for risk-of-bias assessment. This feasibility study is the first to investigate inter-rater reliability of four LLMs applying ROBUST-RCT in comparison to human consensus. Furthermore, it provides data, previously unavailable in the scientific literature, on the inter-rater reliability among different human reviewers applying ROBUST-RCT and among different LLMs.

## METHODS

We designed a feasibility study with humans and LLMs using ROBUST-RCT.

### Sample

We searched for *(“drug therapy”[MeSH Terms]) AND (Randomized Controlled Trial [Publication Type]) AND (“pubmed pmc”[filter])* on PubMed on May 18, 2025, sorted by the platform’s default algorithm. The choice for articles on PubMed Central allows standardization based on the same PDF file. We exported the first 10000 results to a file. We then randomized rows using Google Sheets and selected the first 20 rows in the new order.

### Exclusion Criteria

The full text of each article reporting an RCT was screened for eligibility by independent reviewers. Three reviewers, working in pairs of independent reviewers, screened the full text for exclusion based on predefined criteria (see Supplementary Table 1). Any disagreements on article exclusion were resolved in a consensus discussion.

### Human assessment

ROBUST-RCT proposes itself to be straightforward enough to be used by junior systematic reviewers, with its application for this subset of researchers being part of the tests originally employed to evaluate its usability on the ROBUST-RCT launch paper[13]. Therefore, in our experiment, a group of three early-career researchers, acting as independent reviewers, applied the ROBUST-RCT tool following the ROBUST-RCT manual’s recommendations. The individuals documented their judgments in a standardized spreadsheet.

ROBUST-RCT has 6 items, each with 2 steps, and each step has four possible answers (Supplementary Table 2), except for step 1 of item 6. The value ranges for item 6, step 2 (Supplementary Table 3) followed the thresholds in the example from the ROBUST-RCT manual. Assessment was performed based exclusively on the article. There was an explicit recommendation to disregard other external sources of information—such as study protocols—to preserve the fairness of the assessment conditions. Disagreements between assessors were resolved in a consensus meeting.

To ensure standardization in the application of ROBUST-RCT, only the primary outcome of each study was assessed. Articles that had been excluded by consensus were not subjected to evaluation using ROBUST-RCT. All multiple-choice fields were mandatory.

### AI assessment

A standardized prompt, based on the ROBUST-RCT manual[13], instructed the LLMs to perform the assessment. The input uses the Chain-of-Thought (CoT) strategy[14–16]. Its format includes an initial contextualization, a step-by-step guide on ROBUST-RCT, and an output format. The full CoT prompt is available in Appendix 1.

This prompt was submitted with each study’s article—and, when necessary, supplemental materials. We tested four LLMs: GPT-4-turbo (OpenAI) on May 31, 2025; Gemini 2.5 Pro Preview (Google) on May 30, 2025; DeepSeek-R1 (Hangzhou DeepSeek Artificial Intelligence) on May 31, 2025; and Qwen3-235B-A22B (Alibaba Cloud) on May 31, 2025. The last two models are open-source[17,18]. Given the complexity of the task, advanced reasoning tools were activated when available.

### Statistics

Data from human reviewers were filtered into a final dataset including only those articles submitted for LLM evaluation. The inter-rater reliability analysis between humans and LLMs used only the final consensus ratings from the human reviewers. Statistical analyses were all performed in the R language version 4.4.0. The analysis scripts used irrCAC package[19] and were validated by successfully replicating results from prior papers[4,20] using a publicly available dataset[21].

### Primary Analysis

We established a set of assessments for the main analysis that included all final judgments (step 2 from ROBUST-RCT core items 1 to 6), therefore defined as the “judgment set”. This choice is based on clinical relevance and its contribution to statistical power. From a medical perspective, this set combines the most critical step of the assessments across multiple aspects of RCTs. In data analysis, aggregating these data also enhances precision and stability.

The primary analysis is a Gwet’s AC2 inter-rater reliability coefficient[22] of the judgment set, divided into four distinct analyses comparing human consensus against each of the 4 LLMs. The calculation for this primary analysis used the following parameters: ordinal weights, 4 possible levels, and 95% confidence.

This measurement aims to bring robustness to the inter-rater reliability analysis. Gwet’s AC2 does not suffer from the same issue as Kappa coefficients[23–26], which are strongly affected by the frequency distribution[27], biased in real-world scenarios[28], and may present paradoxical results[29,30]. The choice of parameters, based on those previously used for similar data[20], considers the magnitude of agreement. Thus, distant divergences (e.g., “Definitely Low” and “Definitely High”) are penalized more heavily than close divergences (e.g., “Definitely Low” and “Probably Low”).

This analysis includes a probabilistic benchmark[22] combined with the Landis and Koch classification[31], which considers not only the coefficient but also its standard error (SE). This is necessary due to the differences in precision for the same coefficient (e.g., 0.41) with different SEs (e.g., 0.03 and 0.35), which would limit the interpretation of the results. This problem is mitigated by a process where the probabilities of a coefficient falling into each value range or a higher one are computed by calculating the interval membership probability (IMP). Thus, Gwet’s benchmark allows inferring the minimum classification to which that coefficient belongs with 95% confidence.

### Exploratory Analyses

Three exploratory analyses were defined.

1. Fleiss’ Kappa[32] of the “judgment set” among different human assessors before consensus, using the following parameters: “identity” weight, 4 levels, and 95% confidence. The coefficients were interpreted through the classification from Landis and Koch. This approach aims to provide comparability to previous studies of human inter-rater reliability in risk-of-bias tools.
2. Analysis of the direction of bias for each LLM’s assessments relative to human consensus for the judgment set. To quantify the divergences, numerical values from 0 to 3 were assigned to each of the possible response levels for each ROBUST-RCT item. We then performed a paired Wilcoxon signed-rank test and adjusted p-values with the Bonferroni correction.
3. Assessment of inter-rater reliability using Gwet’s AC2, applied to each multiple-choice step individually, using the same parameters as the primary analysis.

## RESULTS

From our initial sample of 20 randomly selected RCTs, 11 were excluded based on the previously mentioned criteria, resulting in a sample with 9 articles. None of the included studies (Supplementary Table 4) had been previously assessed with ROBUST-RCT. Appendix 2 documents all LLM outputs. Raw data are publicly available[33].

As the judgment set includes step 2 assessments from 6 different core items for 9 RCTs, the primary analysis had a total of 54 comparisons of evaluations between LLMs and human consensus. Step-level exploratory analysis has a smaller sample size (n=9) as it focuses on each multiple-choice step of ROBUST-RCT individually.

### Primary Analysis

The analysis of the judgment set resulted in the following Gwet’s AC2 coefficients (Figure 1) between human consensus and the LLMs (Table 1) and the following benchmarks (Table 2).

**Figure 1.**
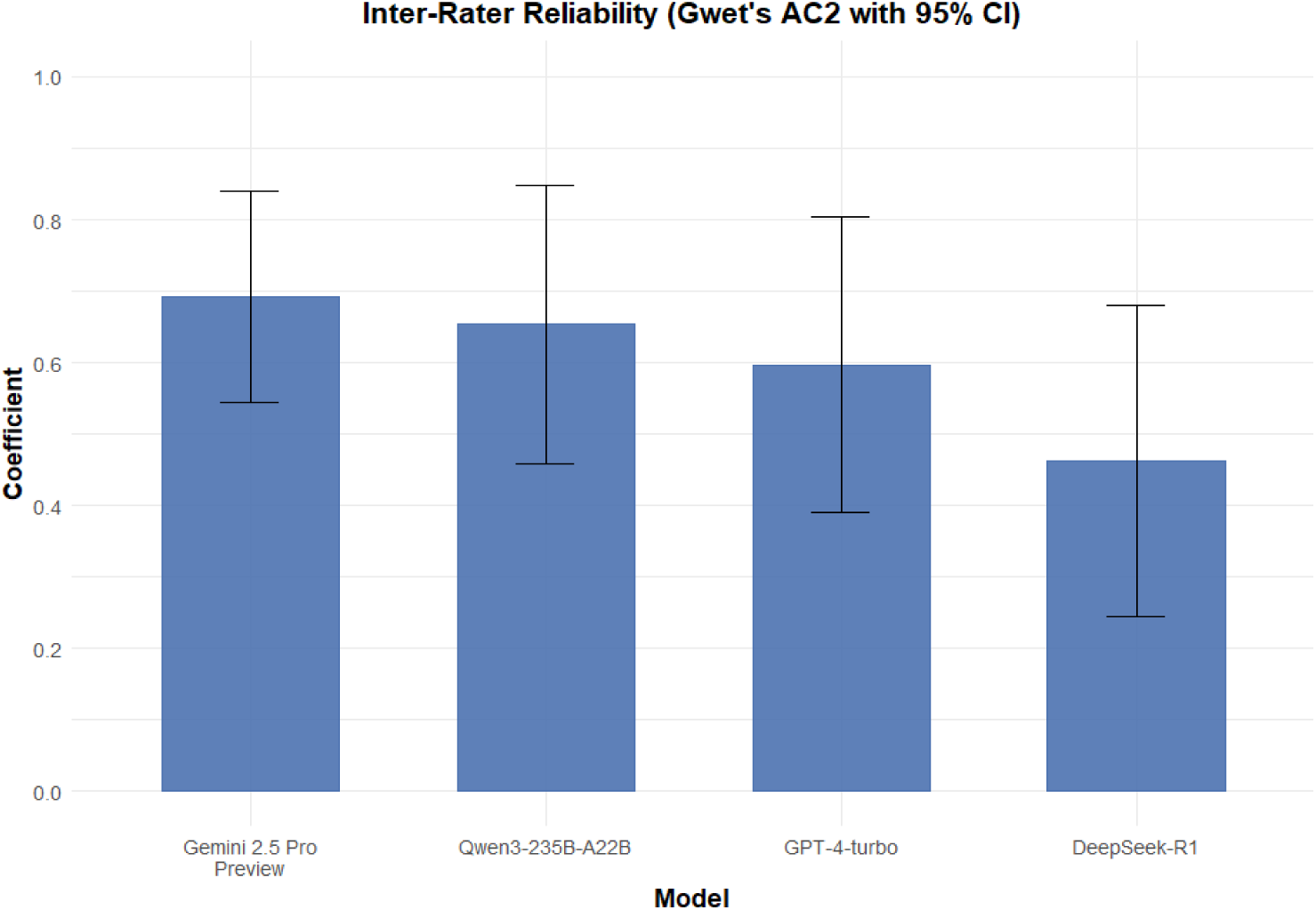
Gwet’s AC2 for the judgment set between human consensus and each LLM.

**Table 1.** Gwet’s AC2 for the judgment set.

| Comparison | Gwet AC2 | Percentage of Agreement | Percentage Expected | Standard Error | 95% Confidence Interval |
| --- | --- | --- | --- | --- | --- |
| GPT-4-turbo | 0.5971 | 0.8364 | 0.3520 | 0.1055 | 0.385, 0.809 |
| Gemini 2.5 Pro Preview | 0.6919 | 0.8703 | 0.3681 | 0.0753 | 0.541, 0.843 |
| DeepSeek-R1 | 0.4621 | 0.7993 | 0.3160 | 0.1108 | 0.24, 0.685 |
| Qwen3-235B- | 0.6532 | 0.8487 | 0.3849 | 0.0991 | 0.454, 0.852 |
| A22B |  |  |  |  |  |

**Table 2.** Benchmark with cumulative probabilities of results in each Landis and Koch classification by Gwet’s method.

| **Landis and Koch** | **Human consensus and GPT-4-turbo** | **Human consensus and Gemini 2.5 Pro Preview** | **Human consensus and DeepSeek-R1** | **Human consensus and Qwen3-235B-A2 2B** |
| --- | --- | --- | --- | --- |
| 0.8 to 1<br>(almost perfect) | 0.02719 | 0.07586 | 0.00115 | 0.06934 |
| 0.6 to 0.8<br>(substantial) | 0.48904 | 0.88868 | 0.1069 | 0.70438 |
| 0.4 to 0.6<br>(moderate) | **0.96912** | **0.99995** | 0.71251 | **0.99467** |
| 0.2 to 0.4<br>(fair) | 0.99992 | 1 | **0.99098** | 1 |
| 0.0 to 0.2<br>(slight) | 1 | 1 | 0.99998 | 1 |
| -1 to 0<br>(poor) | 1 | 1 | 1 | 1 |

The probabilistic benchmarking indicates that, with 95% confidence, the AC2 coefficients fall into the following categories: moderate or higher (human consensus vs. GPT-4-turbo, human consensus vs. Gemini 2.5 Pro Preview, human consensus vs. Qwen3-235B-A22B) and fair or higher (human consensus vs. DeepSeek-R1).

### Exploratory Analyses

The results of the exploratory analyses described in the methods section are reported below.

#### 1. Fleiss’ Kappa of the judgment set (different humans)

The Fleiss’ Kappa was 0.49 (95% CI: 0.30 – 0.6; Supplementary Table 5) between different assessors before consensus, which falls into the “moderate” category of Landis and Koch.

#### 2. Direction of bias analysis (LLMs compared to human consensus)

By assigning ordinal values to the ROBUST-RCT assessments, it was possible to determine whether there was a statistically significant difference between the LLM assessments and human consensus. Mean values and standard deviations are documented in Supplementary Table 6. Wilcoxon signed-rank tests (Supplementary Table 7) revealed significance in the DeepSeek-R1 assessments compared to human consensus (p-value=0.0050; Bonferroni-adjusted p-value=0.0201).

#### 3. Step-level inter-rater reliability (LLMs compared to human consensus)

Given that this specific analysis has a smaller sample size, the confidence interval (CI) crossed the null line at least once for each LLM (Figure 2). It occurred 7 times for GPT-4-turbo, 3 for Gemini 2.5 Pro Preview, 5 for DeepSeek-R1, and 5 for Qwen3-235B-A22B. Appendix 3 provides detailed results and all the benchmarks.

**Figure 2.**
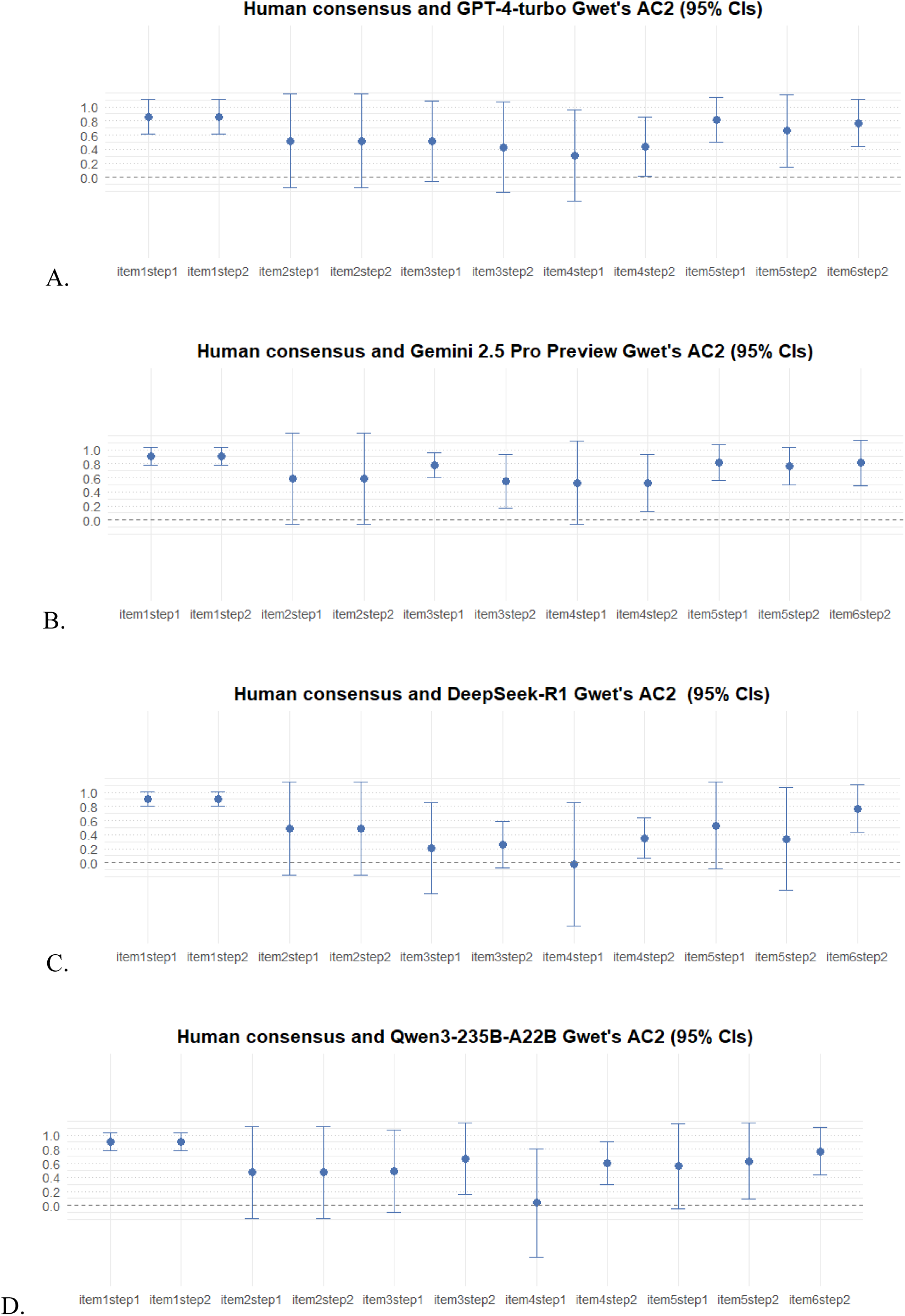
Step-level Gwet’s AC2. A: human consensus and GPT. B: human consensus and Gemini. C: human consensus and Deepseek. D: Human consensus and Qwen.

## DISCUSSION

This study provides the first evidence that LLMs can reliably apply the ROBUST-RCT tool for risk-of-bias assessment. Since three of four tested LLMs showed at least moderate reliability, these findings suggest that integrating these technologies could represent a significant advancement over methods characterized by low reliability and poor usability.

Despite being designed with a small sample, it accomplished the goals we intended for this feasibility study. Our data indicate that it is viable to use LLMs integrated into risk-of-bias assessment processes, a conclusion that had not been answered by previous literature.

Therefore, more than a limitation, this sample size may serve as a basis for further and greater studies. We also believe that its value goes beyond demonstrating a concept, inviting researchers to choose robust approaches for the measure of inter-rater reliability in health sciences.

The roles for LLMs in evidence synthesis are yet to be defined. For the risk-of-bias assessments, it could be used as a tier-breaker, acting as a third reviewer along with two humans. An aggressive method of adoption, however, would be to automate evaluations and then assign human researchers to judge complex cases and disagreements. Semi-automated workflows could be valuable in reducing the burden on research teams, enabling faster updates of living systematic reviews and allowing guideline discussions to be more focused on the findings rather than risk-of-bias assessments.

The strength of our results was tested by rigorous benchmarking and interpretation methods. As demonstrated in our analysis, even a coefficient that might be directly interpreted as “substantial” — such as for Gemini 2.5 Pro Preview (Gwet’s AC2=0.69) — is conservatively classified as “moderate” or higher with 95% confidence. It should also be noted that using other prompt formats, such as Reflection of Thoughts[16], might yield different and potentially better results.

Our inter-rater reliability (0.49 [95% CI: 0.30 – 0.6]) for humans using ROBUST-RCT exceeded previously reported measures of Fleiss’ Kappa for the RoB 2 tool (Figure 3). A prior study[4] found 0.16 (95% CI: 0.08-0.24) for the RoB 2 “overall” domain, classified as “slight”. Another study[2] assessed inter-rater reliability with varying degrees of instruction. Its results for the “overall” judgment ranged from “no agreement” (−0.15) before the use of an implementation document, to “moderate” (0.42) for the last 11 studies evaluated. No CI was reported in that latter study. The employment of junior scientists in our feasibility study instead of seasoned experts also reinforces the usability and consistency of risk-of-bias evaluations with ROBUST-RCT.

**Figure 3.**
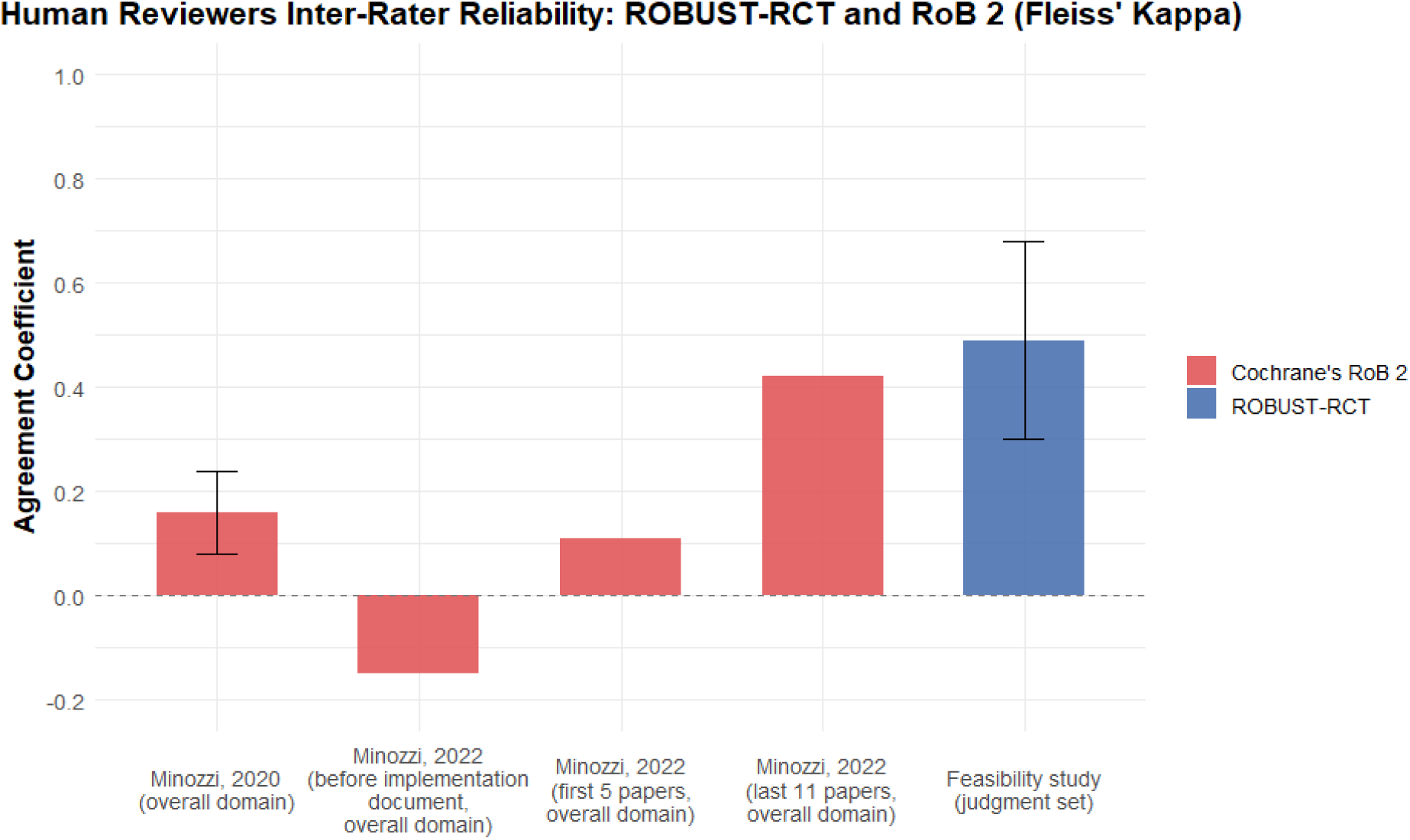
Human reviewers inter-rater reliability, comparing ROBUST-RCT with RoB 2.

Nevertheless, it should be emphasized that a deterministic view — limited to the usual arbitrary Landis and Koch classifications without benchmarking — contrasts with the methodological rigor required for medical sciences. Our work aligns with prior publications[20,22,34], thereby reinforcing the need for robustness in inter-rater coefficient calculation and benchmarking processes in study design.

The direction of bias analysis suggested that the worst-performing LLM — DeepSeek-R1 — may have been more harsh in its evaluations than its human counterparts. DeepSeek-R1’s lack of multimodality may also have contributed to its inferior reliability. While most LLMs have some sort of vision, DeepSeek-R1 just converts submitted PDFs into plain text through optical character recognition. As a result, this process could lead to the loss of information from figures, tables, and diagrams.

Combined with the higher mean value of DeepSeek-R1 (Supplementary Table 6), the Wilcoxon signed-rank test suggests a more stringent evaluation than human consensus. Along with the primary analysis (Gwet’s AC2), this finding suggests that the current version of DeepSeek-R1 may be inadequate due to the presence of alternatives with more consistent assessment results. This result, however, should be interpreted with caution, as our approach assumes human consensus to be the gold standard. It is plausible that, in some instances of disagreement, the AI’s assessment was more accurate.

The last of the exploratory analyses—a Gwet’s AC2 for each multiple-choice step of each item—showed variation in results at the step level, indicating areas for potential improvements in the main study. These data reinforce the plausibility of using LLMs for applying ROBUST-RCT, given the potential margin for prompt improvement. There is also the possibility that human training could give special attention to the aforementioned steps where inter-rater reliability was lower, as human assessment can also be prone to errors. As this analysis used a smaller sample size in comparison to the primary one, its statistical power has been limited.

Just as the possible answers in PROSPERO, the workflow for the assessment of risk of bias in RCTs may quickly change. Our results support a potential role for LLMs to perform evidence synthesis with efficiency, reliability, and objectivity. Improvements in reliability provided by ROBUST-RCT and automation methods could be decisive in reaching a new standard in medical research.

### Limitations

The sample size could limit the precision of the analyses. Focusing on drug interventions could limit the generalization for other interventions (e.g., psychotherapy). The standard algorithm in PubMed search could bias the sampling. The exclusive use of the primary outcome may not reflect the use of ROBUST-RCT in reviews investigating more than one endpoint. LLMs available on web interfaces are subject to updates and modifications by their providers. The level of instruction for the reviewers and the fact that they were not native English speakers could affect inter-rater reliability. We explicitly instructed reviewers to avoid using LLMs and maintain blinding for an independent assessment, but no active barriers for data storage access or website blocking were made. Qualitative analyses were beyond the scope and should be considered in future research.

## Data Availability

Raw data available at Open Science Framework (DOI 10.17605/OSF.IO/CRN64).

https://osf.io/crn64/

## Acknowledgements

In compliance with the statement about generative AI for medical research[35], we transparently report the use of AI technologies. Besides the experiment, our team used Gemini (2.5 Pro Preview, 2.5 Flash, 2.5 Pro) from Google and ChatGPT (GPT-4-turbo, GPT-5) from OpenAI between March 2025 and August 2025 through their web interface. AI applications in research assistance included programming, advising, translating, and text editing. The authors critically evaluated every response and take full responsibility for the integrity of the content in this manuscript. We also thank our colleague and researcher, Sofia Simoni Rossi Fermo, for the proofreading of the English version of the manuscript.

