## Appendix 2 for "Assessment of Bias in Clinical Trials with LLMs Using ROBUST-RCT: A Feasibility Study"

**# GEMINI 2.5 PRO (PREVIEW) OUTPUTS - 30.05.2025 (DDMMYYYY)**

Article: Matsui, 2014

Title: Antibiotic Prophylaxis in Laparoscopic Cholecystectomy: A Randomized Controlled Trial

Journal: PLOS ONE

Outcome: The occurrence of postoperative infections

### **ITEM 1 (Random sequence generation)**

Step 1

Definitely yes

Step 2

Definitely low

Support for judgment for Item 1

The trial explicitly stated the use of an adequate method of generating the random allocation sequence: "Patients were randomized into either of two groups using a computer-generated random number just before the operation..."

### **ITEM 2 (Allocation concealment)**

Step 1

Definitely yes

Step 2

Definitely low

Support for judgment for Item 2

The trial used a clearly satisfactory allocation concealment method: "...by a third party who telephoned the allocation to the operating theatre." This describes central allocation.

### **ITEM 3 (Blinding of participants)**

Step 1

Definitely no

Support for judgment for Item 3, Step 1

The trial did not blind participants. Patients in the Antibiotics group received intravenous antibiotics, while "Patients in the No antibiotics group received no antibiotics." No placebo was mentioned.

Step 2

Probably high

Support for judgment for Item 3, Step 2

Participants were definitely not blinded. Unblinding likely influenced the outcome because participant expectations or behavior (e.g., reporting of symptoms that could lead to investigation and diagnosis of infection, or adherence to other postoperative care) could have been affected by knowledge of their treatment group, potentially influencing the rate of diagnosed postoperative infections.

### **ITEM 4 (Blinding of healthcare providers)**

Step 1

Definitely no

Support for judgment for Item 4, Step 1

The trial explicitly stated that healthcare providers were not blinded: "The surgeons who provided the assigned treatment and interventions were not masked to the patient's group assignment."

Step 2

Probably high

Support for judgment for Item 4, Step 2

Healthcare providers were definitely not blinded. This unblinding likely influenced the outcome, as healthcare provider-initiated co-interventions or their threshold for diagnosing infections could have been affected by knowledge of the patient's allocation, even if protocols for managing suspected infections were in place.

### **ITEM 5 (Blinding of outcome assessors)**

Step 1

Probably yes

Support for judgment for Item 5, Step 1

A key outcome assessor was blinded: "A doctor in charge of the outpatient department who was unaware of the randomization checked the patients' status at least once on postoperative day 8." Many infections, such as wound infections and urinary tract infections, reportedly "developed after discharge" and were thus likely assessed by this blinded doctor. While infections diagnosed during hospitalization (e.g., subhepatic abscesses, colitis ) may have been assessed by unblinded hospital staff (surgeons were unblinded ), the majority of the primary outcome events appear to have been assessed by a blinded individual.

Step 2

Probably low

Support for judgment for Item 5, Step 2

Outcome assessors for a majority of the infection events were probably blinded. Furthermore, many of the postoperative infections involved relatively objective diagnostic criteria (e.g., "pus discharge from the surgical wound", "intra-abdominal abscesses requiring drainage under ultrasonographic control", urinary tract infection diagnosed by a "urine test that microscopically revealed bacteria" ), which would make it less likely that unblinding of some assessors significantly influenced the outcome assessment.

### **ITEM 6 (Outcome data not included in analysis)**

Step 1 (Extract the number of participants who were not included in analysis in each group)

Intervention group → Number of participants not analyzed: 0

Intervention group → Total number of participants: 518

Intervention group → % of participants not analyzed: 0%

Control group → Number of participants not analyzed: 0

Control group → Total number of participants: 519

Control group → % of participants not analyzed: 0%

Overall → Number of participants not analyzed: 0

Overall → Total number of participants: 1037

Overall → % of participants not analyzed: 0%

Step 2

Definitely low

Support for judgment for Item 6, Step 2

The percentage of participants not included in the intention-to-treat analysis for the primary outcome was 0%. This is less than the 5% threshold. The paper states that for the 3 patients who did not visit the hospital after discharge, "By phone two weeks after discharge, these three patients were confirmed not to have any difficulty related to their operation," and the denominators used for the intention-to-treat analysis (518 for Antibiotics, 519 for No antibiotics) match the numbers randomized to each group.

Article: Hauser, 2014

Title: Long-term safety and sustained efficacy of extended-release pramipexole in early and advanced Parkinson's disease

Journal: European Journal of Neurology

Outcome: Unified Parkinson's Disease Rating Scale (UPDRS) Parts II+III score in the preceding double-blind (DB) trials.

### **ITEM 1 (Random sequence generation)**

Step 1

Probably no

Step 2

Probably high

Support for judgment for Item 1

The provided paper describes the preceding studies as "double-blind (DB) trial[s] in which subjects were randomized" but does not provide further details on the method of random sequence generation (e.g., random number table, generator). It also does not mention whether simple, block, or stratified randomization was used, nor does it describe the allocation concealment mechanism within this document. Therefore, it fits the criterion: "Trial described as "randomized" without further details, AND does not meet "Probably Yes/Low" criteria."

### **ITEM 2 (Allocation concealment)**

Step 1

Probably yes

Step 2

Probably low

Support for judgment for Item 2

The preceding studies are described as "double-blind (DB) studies" involving drug treatment (pramipexole ER, pramipexole IR) versus placebo or active comparator, and a "double-dummy design" was used. According to the ROBUST-RCT guidance, for drug trials where participants and healthcare providers were blinded (as implied by "double-blind" and use of placebo/double-dummy), if there is no further information on allocation concealment, a judgment of "Probably Yes / Probably Low" is appropriate. The paper states: "In two double-blind (DB) studies of early PD and one of advanced PD, active-treatment arms received pramipexole immediate release (IR) or extended release (ER), with exposure lasting up to 33 weeks." And, "In all three initial studies, DB treatment was administered t.i.d. - specifically, IR or placebo as morning, afternoon and evening doses, plus placebo or ER as a morning dose (double-dummy design)."

### **ITEM 3 (Blinding of participants)**

Step 1

Probably yes

Support for judgment for Item 3, Step 1

The paper states that the preceding studies were "double-blind (DB)" and involved placebo control or a double-dummy design. For example, "For use in early PD, supporting studies include a 33-week, double-blind (DB) trial [1] in which subjects were randomized to pramipexole ER q.d., pramipexole IR t.i.d. or placebo." "In all three initial studies, DB treatment was administered t.i.d. - specifically, IR or placebo as morning, afternoon and evening doses, plus placebo or ER as a morning dose (double-dummy design)." This indicates that participants were likely blinded. This fits "described as 'single/double/triple blinded' and judgment suggests participants were one of the blinded groups".

Step 2

Probably low

Support for judgment for Item 3, Step 2

Participants were "Probably Yes" blinded in the DB trials. The main outcome, UPDRS Parts II+III, includes Part II which is based on patient reporting of experiences of daily living. Blinding of participants would reduce bias in these self-reports. Given that participants were probably blinded, the risk of bias is "Probably Low".

### **ITEM 4 (Blinding of healthcare providers)**

Step 1

Probably yes

Support for judgment for Item 4, Step 1

The paper describes the preceding studies as "double-blind (DB)" and they involved placebo control or a double-dummy design. For example, "For use in advanced PD, supportive evidence derives from a 33-week DB trial [3] in which subjects...were randomized to the addition of pramipexole ER q.d., pramipexole IR t.i.d. or placebo." The use of "double-blind" suggests healthcare providers were blinded to the treatment allocation. This fits "described as 'single/double/triple blinded' and judgment suggests healthcare providers were one of the blinded groups".

Step 2

Probably low

Support for judgment for Item 4, Step 2

Healthcare providers were "Probably Yes" blinded. Concomitant Parkinson's disease medications were allowed at stable dosages in the DB trials. Blinding of healthcare providers would minimize differential co-interventions based on knowledge of treatment. Therefore, the risk of bias is "Probably Low".

### **ITEM 5 (Blinding of outcome assessors)**

Step 1

Probably yes

Support for judgment for Item 5, Step 1

The preceding trials were "double-blind (DB)". The main outcome was the UPDRS Parts II+III score. Part II is typically patient-reported (and participants were likely blinded, see Item 3). Part III is clinician-assessed. In a double-blind trial, it is expected that the clinician assessing the outcome would also be blinded. The term "double-blind" suggests outcome assessors were part of the blinded groups.

Step 2

Probably low

Support for judgment for Item 5, Step 2

Outcome assessors were "Probably Yes" blinded. While the UPDRS Parts II+III assessment involves some judgment from both the patient (Part II) and the clinician (Part III), if the assessors are blinded, the risk of bias due to unblinding is low. Thus, "Probably Low" is selected because the outcome assessors were probably blinded.

### **ITEM 6 (Outcome data not included in analysis)**

Step 1 (Extract the number of participants who were not included in analysis in each group)

Intervention group → Number of participants not analyzed: Information not provided in this document for the primary analysis of the original DB trials.

Intervention group → Total number of participants: Information not provided in this document for the primary analysis of the original DB trials.

Intervention group → % of participants not analyzed: Information not provided in this document for the primary analysis of the original DB trials.

Control group → Number of participants not analyzed: Information not provided in this document for the primary analysis of the original DB trials.

Control group → Total number of participants: Information not provided in this document for the primary analysis of the original DB trials.

Control group → % of participants not analyzed: Information not provided in this document for the primary analysis of the original DB trials.

Overall → Number of participants not analyzed: Information not provided in this document for the primary analysis of the original DB trials.

Overall → Total number of participants: Information not provided in this document for the primary analysis of the original DB trials.

Overall → % of participants not analyzed: Information not provided in this document for the primary analysis of the original DB trials.

Step 2

Probably high

Support for judgment for Item 6, Step 2

The provided paper (Hauser 2014) describes open-label extensions of previous double-blind (DB) trials. It does not contain specific numbers for participants randomized, or those included/excluded in the primary outcome analysis (UPDRS Parts II+III) of those original DB trials. While Figure 1 shows numbers completing DB trials and entering OL extensions, it does not provide the necessary details (number randomized per arm in DB trials and number analyzed for the primary outcome in DB trials) to calculate the percentage of missing outcome data for the primary analyses of the DB trials themselves. Lack of this information in the provided document prevents a direct assessment based on pre-specified thresholds, and such missing information generally indicates a higher risk of bias or limitations in the reporting being assessed. Therefore, "Probably high" is selected.

Article: Fregni, 2004

Title: Repetitive transcranial magnetic stimulation is as effective as fluoxetine in the treatment of depression in patients with Parkinson's disease

Journal: J Neurol Neurosurg Psychiatry

Outcome: Change in depression scores (Hamilton Rating Scale for Depression - HRSD, and Beck Depression Inventory - BDI)

### **ITEM 1 (Random sequence generation)**

Step 1

Definitely yes

Step 2

Definitely low

Support for judgment for Item 1

The trial explicitly stated the use of an adequate method: "The patients were randomly assigned according to a computer generated randomisation list to one of two groups".

### **ITEM 2 (Allocation concealment)**

Step 1

Probably yes

Step 2

Probably low

Support for judgment for Item 2

The trial involved a placebo drug ("active rTMS and placebo drug treatment" vs "sham rTMS and fluoxetine 20 mg/day") suggesting blinding of participants and healthcare providers for the drug component. For drug trials where participants and healthcare providers were blinded and no further information on allocation concealment is provided, the ROBUST-RCT guidance suggests "Probably Yes / Probably Low". The paper does not provide further specific details on the allocation concealment mechanism (e.g., central allocation, specific envelope procedures) beyond the computer-generated list.

### **ITEM 3 (Blinding of participants)**

Step 1

Probably yes

Support for judgment for Item 3, Step 1

The trial used a placebo drug for Group 1 ("active rTMS and placebo drug treatment") and sham rTMS for Group 2 ("sham rTMS and fluoxetine 20 mg/day"). A "specially designed sham coil was used for sham stimulation". The paper states, "Given that all our patients were naive to rTMS, it is unlikely that they could have guessed the type of treatment." This implies participants were intended to be blinded.

Step 2

Probably low

Support for judgment for Item 3, Step 2

Participants were probably blinded through the use of placebo medication and a sham rTMS procedure. The authors believed the sham was effective for naive patients. While perfect blinding with rTMS can be challenging as "The available sham coils do not produce the same scalp sensation", the effort to use a "specially designed sham coil" and placebo drug makes significant unblinding less likely to have influenced the outcome, or at least, unblinding was unlikely to have markedly influenced the outcome given the measures taken.

### **ITEM 4 (Blinding of healthcare providers)**

Step 1

Probably yes

Support for judgment for Item 4, Step 1

The study is described as "double blind". Group 1 received "active rTMS and placebo drug treatment" and group 2 received "sham rTMS and fluoxetine 20 mg/day". This design, particularly the use of a placebo drug and a sham rTMS coil, suggests that healthcare providers (those administering treatments or managing patient care related to the interventions) were likely intended to be blinded.

Step 2

Probably low

Support for judgment for Item 4, Step 2

Healthcare providers were probably blinded. If blinding was maintained, it is unlikely they would introduce co-interventions that differ systematically between groups. The paper does not detail co-interventions, but the "double blind" design aims to prevent this. Therefore, unblinding was unlikely to have significantly influenced the outcome through provider behavior.

### **ITEM 5 (Blinding of outcome assessors)**

Step 1

Definitely yes

Support for judgment for Item 5, Step 1

The paper explicitly states: "The unified Parkinson's disease rating scale (UPDRS), activities of daily living (ADL), Hamilton rating scale for depression (HRSD), Beck depression inventory (BDI), and mini-mental state examination (MMSE) were assessed by a rater blinded to treatment arm." And further: "The psychiatric and neurological raters, who remained blind to the results of the other raters and to the study group assignment, carried out the evaluations...". This covers the rater-assessed HRSD. The BDI is often self-reported; if so, participant blinding (Item 3) is relevant for BDI. Focusing on HRSD as a key rater-assessed depression outcome, the assessors were blinded.

Step 2

Definitely low

Support for judgment for Item 5, Step 2

Outcome assessors for the Hamilton Rating Scale for Depression (HRSD), a main outcome measure, were definitely blinded as stated: "assessed by a rater blinded to treatment arm" and "The psychiatric and neurological raters... remained blind... to the study group assignment". This meets the criteria for "Definitely Low" risk of bias.

### **ITEM 6 (Outcome data not included in analysis)**

Step 1 (Extract the number of participants who were not included in analysis in each group)

Intervention group (Group 1: rTMS + placebo drug) → Number of participants not analyzed: 1

Intervention group (Group 1: rTMS + placebo drug) → Total number of participants: 21

Intervention group (Group 1: rTMS + placebo drug) → % of participants not analyzed: 4.76%

Control group (Group 2: sham rTMS + fluoxetine) → Number of participants not analyzed: 0

Control group (Group 2: sham rTMS + fluoxetine) → Total number of participants: 21

Control group (Group 2: sham rTMS + fluoxetine) → % of participants not analyzed: 0.00%

Overall → Number of participants not analyzed: 1

Overall → Total number of participants: 42

Overall → % of participants not analyzed: 2.38%

Step 2

Definitely low

Support for judgment for Item 6, Step 2

The paper states: "42 patients were enrolled into two groups", with "group 1: active rTMS and placebo drug treatment (21 patients)" and "group 2: sham rTMS and fluoxetine 20 mg/day (21 patients)". It is also stated: "All patients completed the study, only one patient who had started the study was excluded owing to an unrelated intestinal infection." This means 1 out of 42 (2.38%) participants was not included in the primary analysis of completers. The paper does not explicitly state which group this excluded patient belonged to for the main continuous outcome analyses. To provide per-group numbers as required, the single excluded patient is assigned to Group 1 for this calculation (making its N not analyzed = 1/21, and Group 2 N not analyzed = 0/21); this results in an overall of 1/42 not analyzed. The overall percentage of missing data (1/42 = 2.38%) is less than 5%. Therefore, the risk of bias is "Definitely low".

Article: Scarpone, 2008

Title: The efficacy of prolotherapy for lateral epicondylosis: A pilot study

Journal: Clin J Sport Med

Outcome: resting elbow pain (0-10 Likert scale)

### **ITEM 1 (Random sequence generation)**

Step 1

Definitely yes

Step 2

Definitely low

Support for judgment for Item 1

The trial explicitly stated the use of an adequate method of generating the random allocation sequence: "group assignment was determined by random number table".

### **ITEM 2 (Allocation concealment)**

Step 1

Probably yes

Step 2

Probably low

Support for judgment for Item 2

The trial stated that group assignment was "administered using sealed envelopes". While details on sequential numbering and opacity of the envelopes are not explicitly provided, the study was a double-blind drug trial where participants and the prolotherapist were blinded, and syringes were blinded with an opaque paper sleeve. This aligns with "Probably Yes / Probably Low" criteria: "For drug trials where participants and healthcare providers were blinded: ... stated using envelopes/drug containers but clarity on sequential numbering, opacity, and sealing is lacking."

### **ITEM 3 (Blinding of participants)**

Step 1

Definitely yes

Support for judgment for Item 3, Step 1

The trial explicitly stated that participants were blinded: "Neither prolotherapist nor participant was informed of group status during the study.". The study was also described as a "Double-blind randomized controlled trial".

Step 2

Definitely low

Support for judgment for Item 3, Step 2

Participants were definitely blinded as per the explicit statement in the paper. Although the authors discuss that the prolotherapy solution contained local anesthetics while the control did not, which "may also have affected blinding", they also note this differential effect on post-injection pain "was not observed by the injector". The explicit statement of successful blinding of participants supports a "Definitely Low" risk.

### **ITEM 4 (Blinding of healthcare providers)**

Step 1

Definitely yes

Support for judgment for Item 4, Step 1

The trial explicitly stated that the healthcare provider (prolotherapist) was blinded: "Neither prolotherapist nor participant was informed of group status during the study.". The study was also described as a "Double-blind randomized controlled trial".

Step 2

Definitely low

Support for judgment for Item 4, Step 2

The healthcare provider (prolotherapist) was definitely blinded. Syringes were prepared by the study pharmacist and "blinded with an opaque paper sleeve". While the authors note that "Sodium morrhuate in high concentration has a slightly yellow tint and is slightly more viscous than saline," they conclude that "These issues are unlikely to affect blinding in the setting of pre-filled blinded syringes.". Participants were also discouraged from starting new therapies, attempting to control for co-interventions.

### **ITEM 5 (Blinding of outcome assessors)**

Step 1

Definitely yes

Support for judgment for Item 5, Step 1

The primary outcome was "resting elbow pain, recorded on a 0-10 Likert scale", which is a participant-reported outcome. According to the ROBUST-RCT guidance, "When outcome is participants self-report (e.g., questionnaire), participants are the outcome assessors." As established in Item 3, Step 1, participants were explicitly stated to be blinded.

Step 2

Definitely low

Support for judgment for Item 5, Step 2

Outcome assessors (participants) were definitely blinded. Therefore, the risk of bias related to blinding of outcome assessors is "Definitely Low" based on the guidance "Outcome assessors definitely blinded".

### **ITEM 6 (Outcome data not included in analysis)**

Step 1 (Extract the number of participants who were not included in analysis in each group)

Intervention group → Number of participants not analyzed: 2

Intervention group → Total number of participants: 12

Intervention group → % of participants not analyzed: 16.67%

Control group → Number of participants not analyzed: 2

Control group → Total number of participants: 12

Control group → % of participants not analyzed: 16.67%

Overall → Number of participants not analyzed: 4

Overall → Total number of participants: 24

Overall → % of participants not analyzed: 16.67%

Support for judgment for Item 6, Step 1

The paper states: "24 were consented and randomized to PrT or control injection groups" (N=12 per group). "Two subjects in each group dropped out of the study after baseline data collection but before any injections had been performed.". "Therefore, the analysis of treatment effects included 20 subjects, 10 in each group.". Thus, 2 participants from the PrT group (out of 12) and 2 from the Control group (out of 12) were not included in the analysis of treatment effects. This gives (2/12) * 100 = 16.67% for each group, and (4/24) * 100 = 16.67% overall.

Step 2

Definitely high

Support for judgment for Item 6, Step 2

The percentage of participants not included in the analysis is 16.67%. This meets the threshold for "Definitely high" risk of bias (Missing data ≥ 15%).

Article: Friedman, 2018

Title: A Randomized, Double-Blind, Placebo-Controlled Trial of Naproxen With or Without Orphenadrine or Methocarbamol for Acute Low Back Pain

Journal: Ann Emerg Med

Outcome: Improvement on the Roland-Morris Disability Questionnaire (RMDQ) between ED discharge and one week later

### **ITEM 1 (Random sequence generation)**

Step 1

Definitely yes

Step 2

Definitely low

Support for judgment for Item 1

The trial explicitly stated the use of an adequate method: "The research pharmacist performed randomization in blocks of six based on a sequence generated at http://randomization.com." This indicates the use of a random number generator.

### **ITEM 2 (Allocation concealment)**

Step 1

Definitely yes

Step 2

Definitely low

Support for judgment for Item 2

The trial used a satisfactory allocation concealment method. "The research pharmacist performed randomization...". "Orphenadrine, methocarbamol, and placebo were masked by placing tablets into identical capsules, which were packed with scant amounts of lactose and sealed." Furthermore, "This masking occurred in a secure location inaccessible to ED personnel." This suggests pharmacy-controlled randomization with masked study drugs, ensuring concealment.

### **ITEM 3 (Blinding of participants)**

Step 1

Definitely yes

Support for judgment for Item 3, Step 1

The trial explicitly stated that participants were blinded. It is described as a "randomized, double-blind, comparative effectiveness trial". The study also notes, "Orphenadrine, methocarbamol, and placebo were masked by placing tablets into identical capsules", and "patients did not know if they received active medication or placebo."

Step 2

Definitely low

Support for judgment for Item 3, Step 2

Participants were definitely blinded. The study confirms "patients did not know if they received active medication or placebo." Appendix 3 shows that a considerable number of participants in the placebo group believed they had received a muscle relaxer or were unsure, supporting effective blinding.

### **ITEM 4 (Blinding of healthcare providers)**

Step 1

Definitely yes

Support for judgment for Item 4, Step 1

The trial explicitly stated it was "double-blind". The investigational medications (orphenadrine, methocarbamol, or placebo) were "masked by placing tablets into identical capsules" by a research pharmacist in a "secure location inaccessible to ED personnel". Patients took these medications at home. This setup implies that healthcare providers interacting with the patients regarding the study intervention would be unaware of the treatment allocation.

Step 2

Definitely low

Support for judgment for Item 4, Step 2

Healthcare providers were definitely blinded as per the "double-blind" design and masking procedures for the investigational drugs taken by patients outside the direct supervision of ED staff post-discharge.

### **ITEM 5 (Blinding of outcome assessors)**

Step 1

Definitely yes

Support for judgment for Item 5, Step 1

The primary outcome, improvement on the Roland-Morris Disability Questionnaire (RMDQ), is a patient-reported outcome. As established in Item 3, participants were blinded. The ROBUST-RCT guidance notes: "When outcome is participants self-report (e.g., questionnaire), participants are the outcome assessors." Therefore, the outcome assessors (participants) were blinded. Additionally, "The research associates, who were blinded to assignment, collected all of the data using structured interviews."

Step 2

Definitely low

Support for judgment for Item 5, Step 2

Outcome assessors (participants) were definitely blinded. According to ROBUST-RCT guidance, this leads to a "Definitely Low" risk of bias.

### **ITEM 6 (Outcome data not included in analysis)**

Step 1 (Extract the number of participants who were not included in analysis in each group)

Placebo (Control group) → Number of participants not analyzed: 3

Placebo (Control group) → Total number of participants: 79

Placebo (Control group) → % of participants not analyzed: (3/79) * 100 = 3.8%

Orphenadrine group (Intervention group 1) → Number of participants not analyzed: 2

Orphenadrine group (Intervention group 1) → Total number of participants: 80

Orphenadrine group (Intervention group 1) → % of participants not analyzed: (2/80) * 100 = 2.5%

Methocarbamol group (Intervention group 2) → Number of participants not analyzed: 1

Methocarbamol group (Intervention group 2) → Total number of participants: 81

Methocarbamol group (Intervention group 2) → % of participants not analyzed: (1/81) * 100 = 1.2%

Overall → Number of participants not analyzed: 3 + 2 + 1 = 6

Overall → Total number of participants: 240

Overall → % of participants not analyzed: (6/240) * 100 = 2.5%

Step 2

Definitely low

Support for judgment for Item 6, Step 2

The overall percentage of participants not included in the analysis for the primary outcome is 2.5%. This is less than the pre-specified threshold of < 5% for "Definitely low" risk of bias.

Article: Yang, 2024

Title: COVID-19 Rebound After VV116 vs Nirmatrelvir-Ritonavir Treatment A Randomized Clinical Trial

Journal: JAMA Network Open

Outcome: Viral load rebound (VLR), defined as a half-log increase in viral RNA copies per milliliter compared with treatment completion.

### **ITEM 1 (Random sequence generation)**

Step 1

Definitely yes

Step 2

Definitely low

Support for judgment for Item 1

The trial explicitly stated the use of an adequate method of generating the random allocation sequence. The paper states: "Block randomization with a block size of 4 was used to randomize eligible participants in a 1:1 ratio to receive either VV116 or nirmatrelvir-ritonavir." Block randomization is a recognized adequate method.

### **ITEM 2 (Allocation concealment)**

Step 1

Probably no

Step 2

Probably high

Support for judgment for Item 2

The trial does not provide sufficient detail on the allocation concealment mechanism. While it used block randomization, it also states: "Participants and study staff who were responsible for study drug distribution and recycling were not blinded to the treatment assignment." There is no description of how the allocation sequence was concealed from those enrolling participants or from the unblinded drug distribution staff prior to definitive allocation (e.g., no mention of central randomization, specific pharmacy-controlled procedures ensuring concealment, or sequentially numbered, opaque, sealed envelopes being managed to ensure concealment). This lack of information, coupled with unblinded personnel involved with drug distribution, raises concerns about whether allocation could have been predicted or influenced.

### **ITEM 3 (Blinding of participants)**

Step 1

Definitely no

Support for judgment for Item 3, Step 1

The trial explicitly stated that participants were not blinded. The paper says: "Participants and study staff who were responsible for study drug distribution and recycling were not blinded to the treatment assignment."

Step 2

Probably low

Support for judgment for Item 3, Step 2

Participants were definitely not blinded. The primary outcome, viral load rebound (VLR), is an objective laboratory measurement. While unblinding could lead to differences in co-interventions or participant behavior, it is judged that such factors are "unlikely" to have substantially influenced this objective virological outcome.

### **ITEM 4 (Blinding of healthcare providers)**

Step 1

Definitely yes

Support for judgment for Item 4, Step 1

The trial explicitly stated that healthcare providers involved in treating participants were blinded. The paper states: "Study investigators, including study physicians who treated participants and collected data on medical history and symptoms... were blinded to the treatment assignment."

Step 2

Definitely low

Support for judgment for Item 4, Step 2

Healthcare providers (study physicians who treated participants) were definitely blinded.

### **ITEM 5 (Blinding of outcome assessors)**

Step 1

Definitely yes

Support for judgment for Item 5, Step 1

The trial explicitly stated that outcome assessors for the primary outcome were blinded. The primary outcome is viral load rebound (VLR), which was assessed by laboratory technicians. The paper states: "laboratory technicians who tested for viral load... were blinded to the treatment assignment."

Step 2

Definitely low

Support for judgment for Item 5, Step 2

Outcome assessors (laboratory technicians) were definitely blinded. The primary outcome, VLR, is a laboratory measurement involving minimal judgment in its assessment.

### **ITEM 6 (Outcome data not included in analysis)**

Step 1 (Extract the number of participants who were not included in analysis in each group)

Intervention group → Number of participants not analyzed: 21

Intervention group → Total number of participants: 186

Intervention group → % of participants not analyzed: 11.29%

Control group → Number of participants not analyzed: 8

Control group → Total number of participants: 188

Control group → % of participants not analyzed: 4.26%

Overall → Number of participants not analyzed: 29

Overall → Total number of participants: 374

Overall → % of participants not analyzed: 7.75%

[Data extracted from Figure 1 and text: 374 randomized participants in total. VV116 group: 186 randomized, 165 in full analysis set (FAS). Nirmatrelvir-ritonavir group: 188 randomized, 180 in FAS. The primary outcome was analyzed using the FAS. Thus, not analyzed for VV116 = 186 - 165 = 21. Not analyzed for Nirmatrelvir-ritonavir = 188 - 180 = 8. Overall not analyzed = 374 - (165+180) = 374 - 345 = 29.]

Step 2

Probably low

Support for judgment for Item 6, Step 2

The overall percentage of participants not included in the analysis for the primary outcome is 7.75%. This falls within the pre-specified threshold of "5% ≤ Missing data < 10%", leading to a judgment of "Probably low" risk of bias.

Article: Kabon, 2005

Title: Supplemental Intravenous Crystalloid Administration Does Not Reduce the Risk of Surgical Wound Infection

Journal: Anesth Analg

Outcome: incidence of postoperative wound infections

### **ITEM 1 (Random sequence generation)**

Step 1

Definitely yes

Step 2

Definitely low

Support for judgment for Item 1

"After induction of anesthesia and endotracheal intubation, patients were assigned to small or large perioperative hydration using computer-generated randomized codes that were kept in opaque, sealed, sequentially numbered1 envelopes."

### **ITEM 2 (Allocation concealment)**

Step 1

Definitely yes

Step 2

Definitely low

Support for judgment for Item 2

"After induction of anesthesia and endotracheal intubation, patients were assigned to small or large perioperative hydration using computer-generated randomized codes that were kept in opaque, sealed, sequentially numbered2 envelopes."

### **ITEM 3 (Blinding of participants)**

Step 1

Probably no

Support for judgment for Item 3, Step 1

The study describes that "the surgeons and investigators evaluating wound infections and healing were unable to determine group assignment or perioperative fluid management from patient records", but there is no explicit mention of participants being blinded. Given that the intervention is fluid administration at different rates, it is unlikely participants were blinded.

Step 2

Probably high

Support for judgment for Item 3, Step 2

Participants were likely not blinded. Patient expectations regarding fluid administration could potentially influence self-reported outcomes related to comfort or recovery, although the primary outcome of wound infection is less likely to be directly influenced by participant expectations.

### **ITEM 4 (Blinding of healthcare providers)**

Step 1

Definitely no

Support for judgment for Item 4, Step 1

The paper explicitly states that "the surgeons and investigators evaluating wound infections and healing were unable to determine group assignment or perioperative fluid management from patient records", but does not indicate that healthcare providers administering the fluids were blinded. In fact, different fluid rates were administered, making blinding of healthcare providers responsible for fluid management unlikely.

Step 2

Probably high

Support for judgment for Item 4, Step 2

Healthcare providers responsible for fluid administration were not blinded. This could have influenced co-interventions or aspects of care delivery, potentially affecting patient outcomes.

### **ITEM 5 (Blinding of outcome assessors)**

Step 1

Definitely yes

Support for judgment for Item 5, Step 1

"Surgical wounds were evaluated daily by a physician blinded to group assignment." "After discharge, the same-blinded physician evaluated patients during their two-week clinic visits." "Patients not returning to the clinic were contacted by phone by an investigator blinded to group assignment and treatment."

Step 2

Probably low

Support for judgment for Item 5, Step 2

Outcome assessors were definitely blinded. Although the diagnosis of surgical wound infection involves some judgment, the primary criteria used were "purulent exudate combined with a culture positive for pathogenic bacteria" and "Center for Disease Control criteria for diagnosis of surgical wound infections", which involve objective and standardized assessments.

### **ITEM 6 (Outcome data not included in analysis)**

Step 1 (Extract the number of participants who were not included in analysis in each group)

Intervention group → Number of participants not analyzed: 2

Intervention group → Total number of participants: 131

Intervention group → % of participants not analyzed: 1.5%

Control group → Number of participants not analyzed: 1

Control group → Total number of participants: 125

Control group → % of participants not analyzed: 0.8%

Overall → Number of participants not analyzed: 3

Overall → Total number of participants: 256

Overall → % of participants not analyzed: 1.2%

Step 2

Definitely low

Support for judgment for Item 6, Step 2

Missing data < 5%.

Article: Beretsos, 2009

Title: "hCG priming" effect in controlled ovarian stimulation through a long protocol

Journal: Reproductive Biology and Endocrinology

Outcome: Pregnancy rate

### **ITEM 1 (Random sequence generation)**

Step 1

Definitely yes

Step 2

Definitely low

Support for judgment for Item 1

The trial explicitly stated an adequate method of generating the random allocation sequence: "To perform randomization a random number generator http:// www.random.org was used." "For each patient, a random number between 1 and 100,000 was generated and the patient was allocated to the corresponding group (Group 1 for odd numbers and Group 2 for even numbers)."

### **ITEM 2 (Allocation concealment)**

Step 1

Probably no

Step 2

Probably high

Support for judgment for Item 2

The trial describes random sequence generation but provides no specific information on the method used to conceal the allocation sequence until assignment. Participants and healthcare providers were likely not blinded (see Items 3 and 4). For unblinded drug trials with no further information on allocation concealment, the judgment is "Probably no / Probably high".

### **ITEM 3 (Blinding of participants)**

Step 1

Probably no

Support for judgment for Item 3, Step 1

The paper does not explicitly state participants were blinded. Group 2 received "a fixed 7 days course of 200 IU/day hCG" as a pre-treatment, while Group 1 (control group) did not receive this or an equivalent placebo pre-treatment. This difference in treatment protocol makes blinding of participants unlikely.

Step 2

Probably high

Support for judgment for Item 3, Step 2

Participants were probably not blinded. Knowledge of receiving an additional or different treatment in a fertility trial could influence participant expectations or behaviors (e.g., adherence, reporting of other factors), which could potentially affect the pregnancy rate outcome. Therefore, unblinding likely influenced the outcome.

### **ITEM 4 (Blinding of healthcare providers)**

Step 1

Probably no

Support for judgment for Item 4, Step 1

The paper does not explicitly state that healthcare providers involved in the overall management and administration of the treatment were blinded. Group 2 received an additional 7-day hCG treatment, which would be known to the prescribing/managing clinicians if no identical placebo pre-treatment was administered to Group 1. While the specialist performing oocyte collection and some ultrasounds was blinded, this doesn't cover all healthcare providers managing the stimulation protocol.

Step 2

Probably high

Support for judgment for Item 4, Step 2

Healthcare providers managing the ovarian stimulation were probably not blinded. This knowledge could have influenced co-interventions or decisions during the cycle, such as adjustments to rFSH dosage (which was allowed "according to ovarian response after 5 days of rFSH administration" ), even if unintentionally. Such influences could affect the pregnancy rate.

### **ITEM 5 (Blinding of outcome assessors)**

Step 1

Probably yes

Support for judgment for Item 5, Step 1

The main outcome, clinical pregnancy, was "confirmed by a gestational sac with fetal heartbeat movement seen on transvaginal ultrasound scan". The paper states that "Assessment of follicular growth and endometrial thickness by ultrasound scan as well as oocyte collection were performed by the same fertility specialist being unaware of the study group in which the patient was assigned." It is probable that the same blinded specialist, or another similarly blinded specialist, performed the ultrasound for pregnancy confirmation.

Step 2

Probably low

Support for judgment for Item 5, Step 2

Outcome assessors were probably blinded. The assessment of pregnancy via ultrasound (presence of gestational sac with fetal heartbeat) is a relatively objective measure with minimal judgment, making it unlikely that unblinding would have significantly influenced the assessment.

### **ITEM 6 (Outcome data not included in analysis)**

Step 1 (Extract the number of participants who were not included in analysis in each group)

Intervention group (Group 2) → Number of participants not analyzed: 3

Intervention group (Group 2) → Total number of participants: 22

Intervention group (Group 2) → % of participants not analyzed: 13.64%

Control group (Group 1) → Number of participants not analyzed: 1

Control group (Group 1) → Total number of participants: 28

Control group (Group 1) → % of participants not analyzed: 3.57%

Overall → Number of participants not analyzed: 4

Overall → Total number of participants: 50

Overall → % of participants not analyzed: 8.00%

Step 2

Probably low

Support for judgment for Item 6, Step 2

The overall percentage of participants not included in the analysis for the pregnancy rate outcome is 8.00%. This falls within the 5% ≤ Missing data < 10% threshold, leading to a judgment of "Probably low" risk of bias.

Article: Smith, 2017

Title: A phase I trial of PRN1008, a novel reversible covalent inhibitor of Bruton's tyrosine kinase, in healthy volunteers1

Journal: British Journal of Clinical Pharmacology

Outcome: Safety and tolerability (adverse events)

### **ITEM 1 (Random sequence generation)**

Step 1

Probably yes

Step 2

Probably low

Support for judgment for Item 1

The trial is described as "a two-part randomized, placebo controlled study" and "Both Part A and Part B were designed as double blind, placebo controlled, randomized, ascending dose studies". While the specific method for generating the random sequence (e.g., random number table) is not detailed, the description as "randomized" along with the mention of "double blind" (implying allocation concealment efforts) fits the "Probably yes / Probably low" criteria: "Trial described as 'randomized' without further details, AND...described allocation concealment".

### **ITEM 2 (Allocation concealment)**

Step 1

Probably yes

Step 2

Probably low

Support for judgment for Item 2

The study is described as a "double blind, placebo controlled, randomized" drug trial. According to the ROBUST-RCT guidance, for drug trials where participants and healthcare providers are blinded (implied by "double blind"), if there is "no further information on allocation concealment", the judgment is "Probably yes / Probably low". The paper does not provide specific details on the allocation concealment mechanism (e.g., central randomization, specific envelope procedures).

### **ITEM 3 (Blinding of participants)**

Step 1

Definitely yes

Support for judgment for Item 3, Step 1

The trial is explicitly described as a "double blind, placebo controlled" study. The term "double blind" indicates that participants were blinded to the intervention they received.

Step 2

Definitely low

Support for judgment for Item 3, Step 2

Participants were definitely blinded as the study was "double blind". Therefore, the risk of bias related to participant unblinding influencing the outcome (reporting of adverse events) is definitely low.

### **ITEM 4 (Blinding of healthcare providers)**

Step 1

Definitely yes

Support for judgment for Item 4, Step 1

The trial is explicitly described as a "double blind, placebo controlled" study. The term "double blind" indicates that healthcare providers were blinded to the intervention participants received.

Step 2

Definitely low

Support for judgment for Item 4, Step 2

Healthcare providers were definitely blinded as the study was "double blind". This minimizes the risk that their knowledge of the treatment would influence co-interventions or care, thus the risk of bias is definitely low.

### **ITEM 5 (Blinding of outcome assessors)**

Step 1

Definitely yes

Support for judgment for Item 5, Step 1

The trial is explicitly described as a "double blind, placebo controlled" study. The term "double blind" indicates that outcome assessors (those assessing adverse events) were blinded to the intervention. Adverse events were "categorized using MedDRA, graded using CTCAE criteria and relationship to study drug administration", assessments that could be influenced by lack of blinding.

Step 2

Definitely low

Support for judgment for Item 5, Step 2

Outcome assessors were definitely blinded as the study was "double blind". Although assessment of adverse events (including grading and relationship to study drug ) involves some judgment, the blinding of assessors makes the risk of bias definitely low.

### **ITEM 6 (Outcome data not included in analysis)**

Step 1 (Extract the number of participants who were not included in analysis in each group)

Intervention group → Number of participants not analyzed: 0

Intervention group → Total number of participants: 62

Intervention group → % of participants not analyzed: 0%

Control group → Number of participants not analyzed: 0

Control group → Total number of participants: 18

Control group → % of participants not analyzed: 0%

Overall → Number of participants not analyzed: 0

Overall → Total number of participants: 80

Overall → % of participants not analyzed: 0%

Step 2

Definitely low

Support for judgment for Item 6, Step 2

The paper states, "All subjects completed the study, and there were no withdrawals or dose interruptions." and "Data from all 80 enrolled subjects who received study drug (either PRN1008 or placebo) were included in the safety population." Therefore, 0% of participants had missing outcome data. The pre-specified threshold for "Definitely low" is Missing data < 5%.

**# DEEPTHINK R1 OUTPUTS - 31.05.2025 (DDMMYYYY)**

Article: Matsui, 2014

Title: Antibiotic Prophylaxis in Laparoscopic Cholecystectomy: A Randomized Controlled Trial

Journal: PLoS ONE

Outcome: Overall postoperative infectious complications

\### ITEM 1 (Random sequence generation)

> Step 1

**Probably yes**

> Step 2

**Probably low**

> Support for judgment for Item 1

"Patients were randomized into either of two groups using a computer-generated random number" (Page 2, Methods: Randomization).

---

\### ITEM 2 (Allocation concealment)

> Step 1

**Definitely yes**

> Step 2

**Definitely low**

> Support for judgment for Item 2

"a third party who telephoned the allocation to the operating theatre" (Page 2, Methods: Randomization).

---

\### ITEM 3 (Blinding of participants)

> Step 1

**Definitely no**

> Support for judgment for Item 3, Step 1

"The surgeons who provided the assigned treatment and interventions were not masked to the patient’s group assignment" (Page 2, Methods: Randomization). No placebo or blinding described.

> Step 2

**Probably high**

> Support for judgment for Item 3, Step 2

Unblinding likely influenced co-interventions (e.g., extra antibiotics for fever in No-antibiotics group; Page 4, Methods). Participant expectations could affect symptom reporting or care-seeking for infections.

---

\### ITEM 4 (Blinding of healthcare providers)

> Step 1

**Definitely no**

> Support for judgment for Item 4, Step 1

"The surgeons... were not masked to the patient’s group assignment" (Page 2, Methods: Randomization).

> Step 2

**Probably high**

> Support for judgment for Item 4, Step 2

Unblinding likely influenced antibiotic administration for fever/complications (Page 4, Methods) and discharge decisions, affecting outcome.

---

\### ITEM 5 (Blinding of outcome assessors)

> Step 1

**Definitely yes**

> Support for judgment for Item 5, Step 1

"A doctor in charge of the outpatient department who was unaware of the randomization checked the patients’ status" (Page 2, Methods: Outcome Measures).

> Step 2

**Probably low**

> Support for judgment for Item 5, Step 2

Objective infection criteria (e.g., pus drainage, abscesses, urine tests; Page 3) minimized judgment bias. Blinding further reduced risk.

---

\### ITEM 6 (Outcome data not included in analysis)

> Step 1

Intervention group (Antibiotics) → Number not analyzed: 14 (excluded after conversion to open surgery; Fig. 1)

Intervention group → Total participants: 518

Intervention group → % not analyzed: 2.7%

Control group (No antibiotics) → Number not analyzed: 14 (excluded after conversion to open surgery; Fig. 1)

Control group → Total participants: 519

Control group → % not analyzed: 2.7%

Overall → Number not analyzed: 28

Overall → Total participants: 1037

Overall → % not analyzed: 2.7%

> Step 2

**Definitely low**

> Support for judgment for Item 6, Step 2

Missing data (2.7%) is below the 5% threshold for "Definitely low."

Article: Hauser, 2014

Title: Long-term safety and sustained efficacy of extended-release pramipexole in early and advanced Parkinson's disease

Journal: European Journal of Neurology

Outcome: Change in Unified Parkinson’s Disease Rating Scale (UPDRS) Parts II + III score

\### ITEM 1 (Random sequence generation)

> Step 1

Probably yes

> Step 2

Probably low

> Support for judgment for Item 1

The preceding double-blind (DB) trials described randomization (e.g., "subjects were randomized" in Introduction) but lacked sequence generation details. Allocation concealment was achieved via double-dummy design (Page 2: "DB treatment was administered t.i.d. – specifically, IR or placebo as morning, afternoon and evening doses, plus placebo or ER as a morning dose").

\### ITEM 2 (Allocation concealment)

> Step 1

Probably yes

> Step 2

Probably low

> Support for judgment for Item 2

The trial used a double-dummy design (Page 2: "double-dummy design"), which implies adequate concealment. For drug trials with blinding, this meets "Probably yes" criteria despite no explicit mention of sequential numbering/opacity.

\### ITEM 3 (Blinding of participants)

> Step 1

Definitely no

> Support for judgment for Item 3, Step 1

The extension study was explicitly open-label (Page 2: "open-label (OL) extension study" and "OL treatment was pramipexole ER").

> Step 2

Probably high

> Support for judgment for Item 3, Step 2

Participants were unblinded, and UPDRS Part II includes patient-reported symptoms (e.g., activities of daily living). Participant expectations could influence self-reported outcomes.

\### ITEM 4 (Blinding of healthcare providers)

> Step 1

Definitely no

> Support for judgment for Item 4, Step 1

The extension study was open-label (Page 2: "open-label"), so healthcare providers were aware of treatment.

> Step 2

Probably high

> Support for judgment for Item 4, Step 2

Unblinding could lead to differential co-interventions (e.g., levodopa adjustments). Though concomitant medications were "allowed at stable dosages" (Page 3), dissimilarity between groups was not documented.

\### ITEM 5 (Blinding of outcome assessors)

> Step 1

Probably no

> Support for judgment for Item 5, Step 1

No explicit mention of outcome assessor blinding in the OL extension. The open-label design suggests assessors were likely unblinded.

> Step 2

Definitely high

> Support for judgment for Item 5, Step 2

UPDRS Part III requires clinician judgment during motor examinations. Unblinding could introduce bias in scoring, especially for nuanced assessments like tremor or rigidity.

\### ITEM 6 (Outcome data not included in analysis)

> Step 1

**Early PD (33-week DB trial [1]):**

- Intervention group → Number not analyzed: 77 (368 enrolled - 291 completed)

- Total: 368 → % not analyzed: 20.9%

**Early PD (11–13-week DB trial [2]):**

- Intervention group → Number not analyzed: 26 (143 enrolled - 117 completed)

- Total: 143 → % not analyzed: 18.2%

**Advanced PD (DB trial [3]):**

- Intervention group → Number not analyzed: 62 (391 enrolled - 329 completed)

- Total: 391 → % not analyzed: 15.9%

**Overall:**

- Number not analyzed: 165 (902 enrolled - 737 completed)

- Total: 902 → % not analyzed: 18.3%

> Step 2

Definitely high

> Support for judgment for Item 6, Step 2

Overall missing data (18.3%) exceeds the 15% threshold for "Definitely high" risk. Attrition was similar across groups.

---

**Note:** Disposition data extracted from Page 3 (Fig. 1 and text):

- Early PD [1]: 368 entered OL, 291 completed → 77 not analyzed.

- Early PD [2]: 143 entered OL, 117 completed → 26 not analyzed.

- Advanced PD [3]: 391 entered OL, 329 completed → 62 not analyzed.

Total enrolled: 368 + 143 + 391 = 902. Total completed: 291 + 117 + 329 = 737 → Total not analyzed: 902 - 737 = 165.

Article: Fregni, 2004

Title: Repetitive transcranial magnetic stimulation is as effective as fluoxetine in the treatment of depression in patients with Parkinson’s disease

Journal: Journal of Neurology, Neurosurgery & Psychiatry

Outcome: Depression severity (HRSD and BDI scores)

\### ITEM 1 (Random sequence generation)

> Step 1

**Definitely yes**

> Step 2

**Definitely low**

> Support for judgment for Item 1

"The patients were randomly assigned according to a computer generated randomisation list" (Page 2).

\### ITEM 2 (Allocation concealment)

> Step 1

**Probably no**

> Step 2

**Probably high**

> Support for judgment for Item 2

No mention of allocation concealment methods. Described as "randomised" without details on concealment.

\### ITEM 3 (Blinding of participants)

> Step 1

**Probably yes**

> Support for judgment for Item 3, Step 1

Sham rTMS coil and placebo pills were used. "A specially designed sham coil was used for sham stimulation" and pills were "fluoxetine 20 mg and glucose (placebo)" (Page 1, Page 2).

> Step 2

**Probably high**

> Support for judgment for Item 3, Step 2

Unblinding likely influenced outcome: Psychiatric evaluators guessed treatment correctly in 56% of cases (Page 2). Depression outcomes (HRSD/BDI) are subjective and susceptible to participant expectations.

\### ITEM 4 (Blinding of healthcare providers)

> Step 1

**Probably no**

> Support for judgment for Item 4, Step 1

No explicit statement about healthcare provider blinding. Non-drug trial (rTMS vs. fluoxetine).

> Step 2

**Probably high**

> Support for judgment for Item 4, Step 2

Healthcare providers probably not blinded. Co-interventions (e.g., adjustments in Parkinson’s medications) could influence depression outcomes.

\### ITEM 5 (Blinding of outcome assessors)

> Step 1

**Definitely yes**

> Support for judgment for Item 5, Step 1

"Assessed by a rater blinded to treatment arm" (Page 1).

> Step 2

**Probably high**

> Support for judgment for Item 5, Step 2

Outcome assessors were blinded, but depression scales (HRSD/BDI) require clinical judgment. Unblinding likely influenced assessment: Raters guessed treatment correctly in 56–71% of cases (Page 2).

\### ITEM 6 (Outcome data not included in analysis)

> Step 1

Intervention group (rTMS) → Number not analyzed: 0

Intervention group → Total participants: 21

Intervention group → % not analyzed: 0%

Control group (fluoxetine) → Number not analyzed: 1 (due to intestinal infection)

Control group → Total participants: 21

Control group → % not analyzed: 4.76%

Overall → Number not analyzed: 1

Overall → Total participants: 42

Overall → % not analyzed: 2.38%

> Step 2

**Definitely low**

> Support for judgment for Item 6, Step 2

Overall missing data = 2.38% (<5% threshold).

Article: Scarpone, 2008

Title: The efficacy of prolotherapy for lateral epicondylosis: A pilot study

Journal: Clinical Journal of Sport Medicine

Outcome: Resting elbow pain (0–10 Likert scale)

\### ITEM 1 (Random sequence generation)

> Step 1

Definitely yes

> Step 2

Definitely low

> Support for judgment for Item 1

"The 1:1 randomization scheme was prepared by the lead pharmacist [...] group assignment was determined by random number table."

\### ITEM 2 (Allocation concealment)

> Step 1

Probably yes

> Step 2

Probably low

> Support for judgment for Item 2

"Group assignment was [...] administered using sealed envelopes." (No explicit mention of sequential numbering/opacity).

\### ITEM 3 (Blinding of participants)

> Step 1

Probably no

> Support for judgment for Item 3, Step 1

Non-drug trial; solutions were clear but sodium morrhuate had "slightly yellow tint" and greater viscosity, potentially detectable.

> Step 2

Probably high

> Support for judgment for Item 3, Step 2

Unblinding likely influenced pain reporting (participant self-report outcome); viscosity/tint differences could reveal group allocation.

\### ITEM 4 (Blinding of healthcare providers)

> Step 1

Probably no

> Support for judgment for Item 4, Step 1

Injector (MS) blinded via opaque sleeves, but physical properties (tint/viscosity) might allow detection; non-drug trial.

> Step 2

Probably high

> Support for judgment for Item 4, Step 2

Co-interventions (e.g., NSAID use) were discouraged but not documented; unblinding could affect provider advice/co-interventions.

\### ITEM 5 (Blinding of outcome assessors)

> Step 1

Definitely no

> Support for judgment for Item 5, Step 1

Primary outcome (pain) was self-reported; participants were the outcome assessors and were not blinded (per Item 3).

> Step 2

Definitely high

> Support for judgment for Item 5, Step 2

Outcome involved subjective symptom scoring (Likert scale); unblinding very likely influenced self-reported pain assessments.

\### ITEM 6 (Outcome data not included in analysis)

> Step 1

Intervention group → Number of participants not analyzed: 2

Intervention group → Total number of participants: 12

Intervention group → % of participants not analyzed: 16.7%

Control group → Number of participants not analyzed: 2

Control group → Total number of participants: 12

Control group → % of participants not analyzed: 16.7%

Overall → Number of participants not analyzed: 4

Overall → Total number of participants: 24

Overall → % of participants not analyzed: 16.7%

> Step 2

Definitely high

> Support for judgment for Item 6, Step 2

Overall missing data (16.7%) ≥ 15% (pre-specified threshold for "Definitely high").

\### ROBUST-RCT Assessment

**Article**: Friedman, 2018

**Title**: A Randomized, Double-Blind, Placebo-Controlled Trial of Naproxen With or Without Orphenadrine or Methocarbamol for Acute Low Back Pain

**Journal**: *Annals of Emergency Medicine*

**Outcome**: Improvement in Roland-Morris Disability Questionnaire (RMDQ) between ED discharge and 1 week.

---

\#### **ITEM 1 (Random sequence generation)**

> **Step 1**

Definitely yes

> **Step 2**

Definitely low

> **Support for judgment for Item 1**

"*The research pharmacist performed randomization in blocks of six based on a sequence generated at http://randomization.com.*" (Page 4). This describes an adequate method (online sequence generator).

---

\#### **ITEM 2 (Allocation concealment)**

> **Step 1**

Probably yes

> **Step 2**

Probably low

> **Support for judgment for Item 2**

"*Orphenadrine, methocarbamol, and placebo were masked by placing tablets into identical capsules... packed with scant amounts of lactose and sealed. This masking occurred in a secure location inaccessible to ED personnel.*" (Page 4). Blinded drug containers were used, but sequential numbering/opacity details are unspecified.

---

\#### **ITEM 3 (Blinding of participants)**

> **Step 1**

Probably yes

> **Support for judgment for Item 3, Step 1**

"*Naproxen was not masked. Orphenadrine, methocarbamol, and placebo were masked by placing tablets into identical capsules...*" (Page 4). Participants were blinded to the add-on therapy (placebo-controlled) but aware of naproxen.

> **Step 2**

Probably high

> **Support for judgment for Item 3, Step 2**

The outcome (RMDQ) is a self-reported functional measure. Unblinding to naproxen could influence expectations/co-interventions (e.g., differential use of rescue analgesics). Appendix 4 lists participants attributing outcomes to perceived placebo/ineffectiveness.

---

\#### **ITEM 4 (Blinding of healthcare providers)**

> **Step 1**

Probably yes

> **Support for judgment for Item 4, Step 1**

"*ED personnel*" were denied access to the masking process (Page 4), implying providers were blinded to add-on therapy.

> **Step 2**

Probably high

> **Support for judgment for Item 4, Step 2**

Co-interventions (e.g., referrals, additional prescriptions) could influence outcomes. Providers knew naproxen was given, potentially leading to differential co-interventions (e.g., if perceived as insufficient).

---

\#### **ITEM 5 (Blinding of outcome assessors)**

> **Step 1**

Definitely yes

> **Support for judgment for Item 5, Step 1**

"*The research associates, who were blinded to assignment, collected all of the data using structured interviews.*" (Page 6).

> **Step 2**

Definitely low

> **Support for judgment for Item 5, Step 2**

Outcome assessors were explicitly blinded, and RMDQ is administered via standardized script, minimizing assessment bias.

---

\#### **ITEM 6 (Outcome data not included in analysis)**

> **Step 1**

Intervention group 1 (Naproxen + Orphenadrine) → Number not analyzed: 2 → Total: 80 → %: 2.5%

Intervention group 2 (Naproxen + Methocarbamol) → Number not analyzed: 1 → Total: 81 → %: 1.2%

Control group (Naproxen + Placebo) → Number not analyzed: 3 → Total: 79 → %: 3.8%

Overall → Number not analyzed: 6 → Total: 240 → %: 2.5%

> **Step 2**

Definitely low

> **Support for judgment for Item 6, Step 2**

Overall missing data (2.5%) is below the 5% threshold for "Definitely low" risk.

---

Article: Yang, 2024

Title: COVID-19 Rebound After VV116 vs Nirmatrelvir-Ritonavir Treatment

Journal: JAMA Network Open

Outcome: Viral load rebound (VLR)

\### ITEM 1 (Random sequence generation)

> Step 1

Definitely yes

> Step 2

Definitely low

> Support for judgment for Item 1

"Block randomization with a block size of 4 was used to randomize eligible participants in a 1:1 ratio" (Page 3, "Randomization" section).

\### ITEM 2 (Allocation concealment)

> Step 1

Probably no

> Step 2

Probably high

> Support for judgment for Item 2

No details on allocation concealment methods provided. The trial is unblinded ("Participants and study staff responsible for drug distribution were not blinded") (Page 3, "Randomization" section).

\### ITEM 3 (Blinding of participants)

> Step 1

Definitely no

> Support for judgment for Item 3, Step 1

"Participants and study staff who were responsible for study drug distribution [...] were not blinded" (Page 3, "Randomization" section).

> Step 2

Probably low

> Support for judgment for Item 3, Step 2

Viral load rebound is laboratory-confirmed (RT-qPCR/ddPCR), minimizing influence of participant expectations or co-interventions.

\### ITEM 4 (Blinding of healthcare providers)

> Step 1

Definitely no

> Support for judgment for Item 4, Step 1

"Study investigators, including study physicians who treated participants [...] were blinded" but healthcare providers (implied in "treating physicians") were not explicitly blinded, and drug distribution staff were unblinded (Page 3).

> Step 2

Probably high

> Support for judgment for Item 4, Step 2

Co-interventions (e.g., additional antivirals) could influence viral rebound, and dissimilarity was documented (e.g., 3 participants took azvudine; Page 5, Figure 1).

\### ITEM 5 (Blinding of outcome assessors)

> Step 1

Definitely yes

> Support for judgment for Item 5, Step 1

"Laboratory technicians who tested for viral load [...] were blinded to the treatment assignment" (Page 3, "Randomization" section).

> Step 2

Definitely low

> Support for judgment for Item 5, Step 2

Viral load was measured via RT-qPCR/ddPCR—objective laboratory methods requiring minimal judgment.

\### ITEM 6 (Outcome data not included in analysis)

> Step 1

Intervention group (VV116) → Number not analyzed: 21

Intervention group (VV116) → Total participants: 186

Intervention group (VV116) → % not analyzed: 11.3%

Control group (Nirmatrelvir-ritonavir) → Number not analyzed: 8

Control group (Nirmatrelvir-ritonavir) → Total participants: 188

Control group (Nirmatrelvir-ritonavir) → % not analyzed: 4.3%

Overall → Number not analyzed: 29

Overall → Total participants: 374

Overall → % not analyzed: 7.8%

> Step 2

Probably low

> Support for judgment for Item 6, Step 2

Overall missing data (7.8%) falls within the "Probably low" threshold (5% ≤ missing data < 10%).

---

*Judgments based exclusively on the provided manuscript text and ROBUST-RCT criteria.*

Article: Kabon, 2005

Title: Supplemental Intravenous Crystalloid Administration Does Not Reduce the Risk of Surgical Wound Infection

Journal: Anesthesia & Analgesia

Outcome: Incidence of postoperative wound infection

\### ITEM 1 (Random sequence generation)

> Step 1

Definitely yes

> Step 2

Definitely low

> Support for judgment for Item 1

"Patients were assigned to small or large perioperative hydration using computer-generated randomized codes" (Page 3).

\### ITEM 2 (Allocation concealment)

> Step 1

Definitely yes

> Step 2

Definitely low

> Support for judgment for Item 2

Randomized codes "were kept in opaque, sealed, sequentially numbered envelopes" (Page 3).

\### ITEM 3 (Blinding of participants)

> Step 1

Probably no

> Support for judgment for Item 3, Step 1

Non-drug trial (IV fluid administration) with no explicit mention of participant blinding.

> Step 2

Probably low

> Support for judgment for Item 3, Step 2

Unblinding unlikely to influence outcome (wound infection diagnosed objectively by blinded physicians; participant expectations/behavior unlikely to affect infection risk).

\### ITEM 4 (Blinding of healthcare providers)

> Step 1

Definitely no

> Support for judgment for Item 4, Step 1

Explicitly unblinded: fluid management differed visibly (rates: 8–10 vs. 16–18 mL·kg⁻¹·h⁻¹), and anesthesia records documented group-specific protocols.

> Step 2

Probably low

> Support for judgment for Item 4, Step 2

Co-interventions (e.g., additional fluids, antiemetics) were protocolized based on objective criteria (e.g., urine output, MAP) and were similar between groups (Table 2: hemodynamics, FiO₂, core temperature). No evidence of differential co-interventions affecting infection.

\### ITEM 5 (Blinding of outcome assessors)

> Step 1

Definitely yes

> Support for judgment for Item 5, Step 1

"Surgical wounds were evaluated daily by a physician blinded to group assignment" and records were sealed to prevent unblinding (Page 4).

> Step 2

Definitely low

> Support for judgment for Item 5, Step 2

Outcome assessors were explicitly blinded, and infection diagnosis involved objective criteria (purulent exudate + culture; CDC criteria).

\### ITEM 6 (Outcome data not included in analysis)

> Step 1

Intervention group (Small volume) → Number of participants not analyzed: 1

Intervention group (Small volume) → Total number of participants: 125

Intervention group (Small volume) → % of participants not analyzed: 0.80%

Control group (Large volume) → Number of participants not analyzed: 2

Control group (Large volume) → Total number of participants: 131

Control group (Large volume) → % of participants not analyzed: 1.53%

Overall → Number of participants not analyzed: 3

Overall → Total number of participants: 256

Overall → % of participants not analyzed: 1.17%

> Step 2

Definitely low

> Support for judgment for Item 6, Step 2

Overall missing data (1.17%) is below the 5% threshold for "Definitely low."

Article: Beretsos, 2009

Title: "hCG priming" effect in controlled ovarian stimulation through a long protocol

Journal: Reproductive Biology and Endocrinology

Outcome: Pregnancy rate

\### ITEM 1 (Random sequence generation)

> Step 1

Definitely yes

> Step 2

Definitely low

> Support for judgment for Item 1

"The patients were randomly assigned into two groups... To perform randomization a random number generator http://www.random.org was used."

\### ITEM 2 (Allocation concealment)

> Step 1

Probably no

> Step 2

Probably high

> Support for judgment for Item 2

No description of allocation concealment methods. The trial is non-drug (hCG vs. no hCG), and no details on envelopes/containers were provided.

\### ITEM 3 (Blinding of participants)

> Step 1

Definitely no

> Support for judgment for Item 3, Step 1

No mention of participant blinding. The intervention (hCG injection) vs. control (no hCG) is inherently unblinded in this non-drug trial.

> Step 2

Probably high

> Support for judgment for Item 3, Step 2

Unblinding likely influenced outcome (pregnancy rate) due to potential participant expectations/co-interventions (e.g., stress, lifestyle changes).

\### ITEM 4 (Blinding of healthcare providers)

> Step 1

Probably no

> Support for judgment for Item 4, Step 1

No explicit statement on blinding. Non-drug trial without matched placebos or "double dummy" design.

> Step 2

Probably high

> Support for judgment for Item 4, Step 2

Unblinding likely influenced co-interventions (e.g., dose adjustments, monitoring intensity) affecting pregnancy rates.

\### ITEM 5 (Blinding of outcome assessors)

> Step 1

Definitely yes

> Support for judgment for Item 5, Step 1

"Assessment of follicular growth... performed by the same fertility specialist being unaware of the study group... evaluation of oocyte maturation... by a unique embryologist who was blinded."

> Step 2

Definitely low

> Support for judgment for Item 5, Step 2

Outcome assessors were blinded, minimizing bias in pregnancy confirmation (ultrasound with fetal heartbeat).

\### ITEM 6 (Outcome data not included in analysis)

> Step 1

Intervention group → Number of participants not analyzed: 3

Intervention group → Total number of participants: 22

Intervention group → % of participants not analyzed: 13.64%

Control group → Number of participants not analyzed: 1

Control group → Total number of participants: 28

Control group → % of participants not analyzed: 3.57%

Overall → Number of participants not analyzed: 4

Overall → Total number of participants: 50

Overall → % of participants not analyzed: 8.00%

> Step 2

Probably low

> Support for judgment for Item 6, Step 2

Overall missing data (8%) is ≥5% but <10%, meeting the "Probably low" threshold.

Article: Smith, 2017

Title: A phase I trial of PRN1008, a novel reversible covalent inhibitor of Bruton’s tyrosine kinase, in healthy volunteers

Journal: British Journal of Clinical Pharmacology

Outcome: BTK occupancy in peripheral blood mononuclear cells (PBMCs)

\### ITEM 1 (Random sequence generation)

> Step 1

Probably yes

> Step 2

Probably low

> Support for judgment for Item 1

"The study was conducted in two parts [...] Both Part A and Part B were designed as **double blind, placebo controlled, randomized**, ascending dose studies" (Page 2). The paper describes the trial as "randomized" without specifying the sequence generation method but mentions "randomized" in the context of ascending-dose design and allocation concealment (placebo control).

\### ITEM 2 (Allocation concealment)

> Step 1

Probably yes

> Step 2

Probably low

> Support for judgment for Item 2

No explicit allocation concealment details are provided. However, it is a **blinded drug trial** ("double blind, placebo controlled") with matching placebo, satisfying "Probably Yes/Low" for drug trials where blinding is used.

\### ITEM 3 (Blinding of participants)

> Step 1

Definitely yes

> Support for judgment for Item 3, Step 1

"**Double blind**, placebo controlled" (Page 2). Placebo was administered identically as a liquid formulation, confirming participant blinding.

> Step 2

Definitely low

> Support for judgment for Item 3, Step 2

Participants were blinded, and the outcome (BTK occupancy) is an **objective laboratory measure** unaffected by participant expectations.

\### ITEM 4 (Blinding of healthcare providers)

> Step 1

Definitely yes

> Support for judgment for Item 4, Step 1

"**Double blind**, placebo controlled" (Page 2) implies healthcare providers were blinded.

> Step 2

Definitely low

> Support for judgment for Item 4, Step 2

Healthcare providers were blinded, and co-interventions were unlikely to influence **BTK occupancy** (a biochemical endpoint).

\### ITEM 5 (Blinding of outcome assessors)

> Step 1

Probably yes

> Support for judgment for Item 5, Step 1

No explicit statement, but the **double-blind design** and laboratory-based outcome (BTK occupancy via fluorescence scanning) imply outcome assessors were likely blinded.

> Step 2

Definitely low

> Support for judgment for Item 5, Step 2

BTK occupancy was quantified using **fluorescence scanners and immunoblotting** (Page 3), an objective method with minimal assessment judgment.

\### ITEM 6 (Outcome data not included in analysis)

> Step 1

Intervention group → Number of participants not analyzed: 0

Intervention group → Total number of participants: 62

Intervention group → % of participants not analyzed: 0%

Control group → Number of participants not analyzed: 0

Control group → Total number of participants: 18

Control group → % of participants not analyzed: 0%

Overall → Number of participants not analyzed: 0

Overall → Total number of participants: 80

Overall → % of participants not analyzed: 0%

> Step 2

Definitely low

> Support for judgment for Item 6, Step 2

"All subjects completed the study, and there were no withdrawals or dose interruptions" (Page 3). Missing data = 0% (<5% threshold for "Definitely low").

#GPT4-TURBO (WITH “THINK FOR LONGER” ACTIVATED) OUTPUT [31-05-2025]

Article: Matsui, 2014
 Title: Antibiotic Prophylaxis in Laparoscopic Cholecystectomy: A Randomized Controlled Trial
 Journal: PLoS ONE
 Outcome: Overall postoperative infectious complications

### **ITEM 1 (Random sequence generation)**

Step 1
 “Definitely yes”

Step 2
 “Definitely low”

Support for judgment for Item 1
 “Patients were randomized into either of two groups using a computer-generated random number just before the operation by a third party who telephoned the allocation to the operating theatre” (Methods, Randomization) . This explicit use of a computer‐generated random number constitutes an adequate random sequence generation method, and thus poses a definitely low risk of bias.

### **ITEM 2 (Allocation concealment)**

Step 1
 “Definitely yes”

Step 2
 “Definitely low”

Support for judgment for Item 2
 “Patients were randomized … using a computer-generated random number just before the operation by a third party who telephoned the allocation to the operating theatre” (Methods, Randomization) . The involvement of a third party providing allocation via telephone represents central allocation, a clearly satisfactory concealment mechanism, giving definitely low risk of bias.

### **ITEM 3 (Blinding of participants)**

Step 1
 “Definitely no”

Support for judgment for Item 3, Step 1
 “No masking was performed: ‘The surgeons who provided the assigned treatment and interventions were not masked to the patient’s group assignment’” (Methods, Randomization) . There is no mention of placebo or participant blinding, so participants were definitely not blinded.

Step 2
 “Probably low”

Support for judgment for Item 3, Step 2
 Although participants were definitely not blinded, the primary outcome (postoperative infections) is measured by objective criteria (culture‐confirmed wound infection, drainage of abscess, documented urinary or respiratory infection). Unblinding of participants is unlikely to have influenced the true occurrence of these objectively diagnosed infections; therefore the risk of bias related to participant blinding is probably low.

### **ITEM 4 (Blinding of healthcare providers)**

Step 1
 “Definitely no”

Support for judgment for Item 4, Step 1
 “The surgeons who provided the assigned treatment and interventions were not masked to the patient’s group assignment” (Methods, Randomization) . Hence healthcare providers were explicitly unblinded.

Step 2
 “Probably high”

Support for judgment for Item 4, Step 2
 Healthcare providers were definitely not blinded, and unblinding could plausibly have influenced co-interventions (e.g., closer intraoperative aseptic technique, differential postoperative management or threshold for extra antibiotics), which in turn could have influenced infection rates. Thus unblinding of providers is likely to have affected the outcome and poses a probably high risk of bias.

### **ITEM 5 (Blinding of outcome assessors)**

Step 1
 “Probably yes”

Support for judgment for Item 5, Step 1
 “Examination for SSIs and other infectious diseases were made until hospital discharge and then again at the first postoperative visit. All patients were followed up … by a doctor in charge of the outpatient department who was unaware of the randomization” (Methods, Outcome Measures) . Although not every in-hospital assessor is explicitly stated to be blinded, the outpatient assessor (who determined post‐discharge infections) was unaware of allocation. In addition, this was a drug trial with objective laboratory‐confirmed endpoints, making it probable that outcome assessors were blinded to group assignment.

Step 2
 “Probably low”

Support for judgment for Item 5, Step 2
 Outcome assessment involved objective measures (wound cultures, need for drainage, documented urinary pathogen, etc.), and the doctor performing the postoperative outpatient assessments was unaware of randomization. Consequently, even if some in-hospital assessors may not have been formally blinded, unblinding is unlikely to have influenced these largely objective infection endpoints, leading to a probably low risk of bias.

### **ITEM 6 (Outcome data not included in analysis)**

Step 1 (Extract the number of participants who were not included in analysis in each group)

Intervention group → Number of participants not analyzed: 0
 Intervention group → Total number of participants: 518
 Intervention group → % of participants not analyzed: 0%

Control group → Number of participants not analyzed: 0
 Control group → Total number of participants: 519
 Control group → % of participants not analyzed: 0%

Overall → Number of participants not analyzed: 0
 Overall → Total number of participants: 1,037
 Overall → % of participants not analyzed: 0%

Step 2
 “Definitely low”

Support for judgment for Item 6, Step 2
 All 1,037 randomized participants were included in the intention‐to‐treat analysis (518 in the Antibiotics arm and 519 in the No‐antibiotics arm). No participants were excluded from outcome analysis, yielding 0% missing data, which is well below the 5% threshold for “definitely low” risk of bias.

Article: Hauser, 2014
 Title: Long-term safety and sustained efficacy of extended-release pramipexole in early and advanced Parkinson’s disease
 Journal: European Journal of Neurology
 Outcome: Change in UPDRS Parts II + III score at end of open-label extension (sustained efficacy)

### **ITEM 1 (Random sequence generation)**

Step 1
 “Definitely no”

Step 2
 “Definitely high”

Support for judgment for Item 1
 This paper describes an open-label extension, not a randomized controlled comparison. There is no description of any random allocation procedure for the extension phase: “In both of the extensions, the OL treatment was pramipexole ER, taken q.d. … all subjects entered open-label ER” (Methods section, “Study designs”). Because there is no randomization at this stage, the risk of bias from sequence generation is “Definitely no” and thus “Definitely high.”

### **ITEM 2 (Allocation concealment)**

Step 1
 “Definitely no”

Step 2
 “Definitely high”

Support for judgment for Item 2
 Since all participants in the extension are simply continued on pramipexole ER without any further allocation, there is no allocation process to conceal: “all subjects took ER q.d. for up to 80 weeks” (Methods, Study designs). Because allocation concealment is impossible in a single-arm, open-label extension, we judge “Definitely no” and thus “Definitely high.”

### **ITEM 3 (Blinding of participants)**

Step 1
 “Definitely no”

Support for judgment for Item 3, Step 1
 The extension is explicitly open-label, so participants know they are on pramipexole ER: “This report describes the results of the extensions … In both of the extensions, the OL treatment was pramipexole ER, taken q.d.” (Introduction/Methods). There is no mention of placebo or concealment of assignment. Hence participants were not blinded.

Step 2
 “Definitely high”

Support for judgment for Item 3, Step 2
 Participants were aware of ongoing pramipexole ER throughout; patient expectations and co-interventions could easily influence reported subjective symptoms (e.g., UPDRS Part II or self-reported adverse events). Thus, “Definitely high” risk of bias from lack of participant blinding.

### **ITEM 4 (Blinding of healthcare providers)**

Step 1
 “Definitely no”

Support for judgment for Item 4, Step 1
 No aspect of the extension describes blinding of treating clinicians or study staff; it is open-label throughout: “Subjects from the preceding DB trials … entered the extension and took ER q.d., with dosage adjustment permitted” (Methods, Study designs). Therefore, providers knew treatment assignment.

Step 2
 “Definitely high”

Support for judgment for Item 4, Step 2
 Healthcare providers could alter management (for example, adjusting concomitant PD medications, titrating ER dose) based on knowledge of treatment assignment. This open-label design makes provider-initiated co-interventions likely to differ systematically, thus “Definitely high.”

### **ITEM 5 (Blinding of outcome assessors)**

Step 1
 “Probably no”

Support for judgment for Item 5, Step 1
 There is no statement that UPDRS raters were blinded to previous DB assignment or to the fact that all patients are now on ER. Because the extension staff assessed UPDRS in an unblinded context, but the primary scale (UPDRS Parts II + III) in advanced PD uses “Part III … scored during best ‘on’ time,” which is more objective, we can only say “Probably no” rather than “Definitely no.” However, in early PD, both Parts II (self-reported) and III (clinician scored) are subject to expectation bias if the rater knows the patient is on active treatment.

Step 2
 “Definitely high”

Support for judgment for Item 5, Step 2
 Because outcome assessors were aware that every participant was receiving open-label pramipexole ER, and the main outcome (UPDRS II + III) involves clinician judgment (Part III) and patient report (Part II), unblinding could easily influence scoring. Thus, “Definitely high.”

### **ITEM 6 (Outcome data not included in analysis)**

Step 1 (Extract the number of participants who were not included in analysis in each group)

Although this is an extension (all on active treatment), we treat ex-DB-ER and ex-DB-IR as separate groups for the main outcome (change in UPDRS II + III from the DB baseline). The paper reports:

- “Of 292 early-PD patients from the preceding 33-week DB trial … amongst 234 observed cases, adjusted mean changes …”
   → Ex-DB-ER + Ex-DB-IR combined entering extension: 292
   → Number analyzed at OL end-point: 234
   → Number not analyzed: 292 – 234 = 58
   → % not analyzed: (58/292) × 100 ≈ 19.9%

There is no separate count by arm in the extension publication, so we report combined numbers. (There is no true “control group” in this extension—both arms are now on ER.)

Intervention group → Number of participants not analyzed: 58
 Intervention group → Total number of participants: 292
 Intervention group → % of participants not analyzed: 19.9%

(If one wishes to split by ex-DB-ER vs. ex-DB-IR, the extension paper does not provide separate N for observed cases in each subgroup. Therefore, we use the aggregate.)

Step 2
 “Definitely high”

Support for judgment for Item 6, Step 2
 Overall missing data (19.9%) exceeds the 15% threshold. According to the pre-specified thresholds, missing data ≥15% dictates “Definitely high.”

**Summary of Thresholds Used for Item 6**

- Missing data < 5% → Definitely low
- 5% ≤ Missing data < 10% → Probably low
- 10% ≤ Missing data < 15% → Probably high
- Missing data ≥ 15% → Definitely high

Since 19.9% of participants (58 of 292) were not included in the UPDRS II + III analysis at OL end-point, this falls into “Definitely high.”

Article: Fregni, 2004
 Title: Repetitive transcranial magnetic stimulation is as effective as fluoxetine in the treatment of depression in patients with Parkinson’s disease
 Journal: Journal of Neurology, Neurosurgery & Psychiatry
 Outcome: Change in Hamilton Rating Scale for Depression (HRSD) scores

### **ITEM 1 (Random sequence generation)**

Step 1
 “Definitely yes”

Step 2
 “Definitely low”

Support for judgment for Item 1
 The Methods section states, “The patients were randomly assigned according to a computer generated randomisation list to one of two groups: group 1: active rTMS and placebo drug treatment (21 patients); group 2: sham rTMS and fluoxetine 20 mg/day (21 patients)” (). A computer‐generated list constitutes an adequate method for generating a random allocation sequence, fulfilling the criteria for “Definitely yes,” and thus indicates “Definitely low” risk of bias.

### **ITEM 2 (Allocation concealment)**

Step 1
 “Probably no”

Step 2
 “Probably high”

Support for judgment for Item 2
 No details are provided on how the allocation sequence was concealed (e.g., central allocation or sealed envelopes). The paper merely states that patients “were randomly assigned according to a computer generated randomisation list” without describing how the sequence was implemented or hidden from recruiters and enrolling investigators (). Because there is no mention of any mechanism (such as sequentially numbered, opaque, sealed envelopes), this meets the criteria for “Probably no,” corresponding to “Probably high” risk of bias owing to lack of information on allocation concealment.

### **ITEM 3 (Blinding of participants)**

Step 1
 “Definitely yes”

Support for judgment for Item 3, Step 1
 Participants received either “active rTMS and placebo drug” or “sham rTMS and fluoxetine 20 mg/day,” using “a specially designed sham coil” identical in appearance to the active coil (). This explicit use of a placebo pill and a sham coil ensures that participants were blinded to their treatment assignment.

Step 2
 “Definitely low”

Support for judgment for Item 3, Step 2
 Because participants were definitely blinded via a placebo pill and a sham coil—and given that unblinding is very unlikely to have influenced the main depression outcome measured by HRSD—this satisfies the criterion for “Definitely low” risk of bias related to participant blinding.

### **ITEM 4 (Blinding of healthcare providers)**

Step 1
 “Probably no”

Support for judgment for Item 4, Step 1
 The paper does not report that the clinicians delivering rTMS/sham or dispensing pills were blinded. Although sham coils were used, there is no statement that operators were unaware of coil type, nor is there any description of packaging or masking procedures for fluoxetine versus placebo. In the absence of explicit blinding of providers, this meets the criteria for “Probably no.”

Step 2
 “Probably high”

Support for judgment for Item 4, Step 2
 Because healthcare providers were probably not blinded—and provider knowledge could have influenced co‐interventions such as encouragement or ancillary care—this satisfies the criterion for “Probably high” risk of bias related to provider blinding.

### **ITEM 5 (Blinding of outcome assessors)**

Step 1
 “Definitely yes”

Support for judgment for Item 5, Step 1
 The Methods section states that HRSD, BDI, MMSE, and UPDRS were “assessed by a rater blinded to treatment arm” (). This clearly indicates that outcome assessors were blinded.

Step 2
 “Definitely low”

Support for judgment for Item 5, Step 2
 Because outcome assessors were definitely blinded and the main outcome (HRSD) is a structured rating scale administered by the blinded rater, unblinding could not have influenced depression scoring. This fulfills “Definitely low” risk of bias.

### **ITEM 6 (Outcome data not included in analysis)**

Step 1 (Extract the number of participants who were not included in analysis in each group)

Intervention group → Number of participants not analyzed: 1
 Intervention group → Total number of participants: 21
 Intervention group → % of participants not analyzed: 4.8%

Control group → Number of participants not analyzed: 0
 Control group → Total number of participants: 21
 Control group → % of participants not analyzed: 0%

Overall → Number of participants not analyzed: 1
 Overall → Total number of participants: 42
 Overall → % of participants not analyzed: 2.4%

Step 2
 “Definitely low”

Support for judgment for Item 6, Step 2
 The overall percentage of participants not included in the analysis is 2.4%, which is below the 5% threshold. Thus, according to the pre‐specified thresholds (< 5% missing = “Definitely low”), the risk of bias from missing outcome data is “Definitely low.”

Article: Scarpone, 2008
 Title: The efficacy of prolotherapy for lateral epicondylosis: A pilot study
 Journal: Clinical Journal of Sport Medicine
 Outcome: Resting elbow pain (0–10 Likert scale at 16 weeks)

### **ITEM 1 (Random sequence generation)**

Step 1
 “Definitely yes”

Step 2
 “Definitely low”

Support for judgment for Item 1
 The Methods section states: “The 1:1 randomization scheme was prepared by the lead pharmacist of Trinity Health Systems; group assignment was determined by random number table and administered using sealed envelopes.” This explicitly describes an adequate method (random number table) for generating the allocation sequence, satisfying “Definitely yes,” and indicates a very low likelihood of bias, thus “Definitely low.”

### **ITEM 2 (Allocation concealment)**

Step 1
 “Probably yes”

Step 2
 “Probably low”

Support for judgment for Item 2
 The same sentence in Methods—“group assignment was … administered using sealed envelopes” —suggests use of an envelope-based concealment method. Because the paper does not explicitly state that envelopes were sequentially numbered, opaque, and opened in order, we cannot be certain it meets the “Definitely yes” criteria. However, the description of sealed envelopes prepared by a pharmacist for allocation implies adequate concealment, supporting “Probably yes.” There is no indication of open allocation or quasi-random methods, so risk is probably low.

### **ITEM 3 (Blinding of participants)**

Step 1
 “Definitely yes”

Support for judgment for Item 3, Step 1
 The abstract and Methods both describe this as a “double-blind randomized controlled trial.” Specifically, “Neither prolotherapist nor participant was informed of group status during the study.” That statement explicitly confirms participants were blinded, thus “Definitely yes.”

Step 2
 “Definitely low”

Support for judgment for Item 3, Step 2
 Because participants were definitely blinded, there is very low risk that knowledge of group assignment influenced self-reported pain (the primary outcome). This meets the “Definitely low” criterion.

### **ITEM 4 (Blinding of healthcare providers)**

Step 1
 “Definitely yes”

Support for judgment for Item 4, Step 1
 Again, “Neither prolotherapist nor participant was informed of group status during the study.” This explicitly states that providers (prolotherapist) were blinded, so “Definitely yes.”

Step 2
 “Definitely low”

Support for judgment for Item 4, Step 2
 With providers blinded, it is very unlikely that provider behavior or co-interventions differed systematically by group. Thus, risk related to provider blinding is “Definitely low.”

### **ITEM 5 (Blinding of outcome assessors)**

Step 1
 “Definitely yes”

Support for judgment for Item 5, Step 1
 Since the primary outcome (resting elbow pain) is participant-reported, and participants were blinded (“Neither prolotherapist nor participant was informed of group status”), the outcome assessors (participants) were also blinded.

Step 2
 “Definitely low”

Support for judgment for Item 5, Step 2
 Because outcome assessment (pain rating) was self-reported by blinded participants, there is minimal chance that lack of blinding biased the pain measurement. Therefore, “Definitely low.”

### **ITEM 6 (Outcome data not included in analysis)**

Step 1 (Extract the number of participants who were not included in analysis in each group)
 Intervention group → Number of participants not analyzed: 2
 Intervention group → Total number of participants: 12
 Intervention group → % of participants not analyzed: 16.7%

Control group → Number of participants not analyzed: 2
 Control group → Total number of participants: 12
 Control group → % of participants not analyzed: 16.7%

Overall → Number of participants not analyzed: 4
 Overall → Total number of participants: 24
 Overall → % of participants not analyzed: 16.7%

Step 2
 “Definitely high”

Support for judgment for Item 6, Step 2
 Four randomized subjects (two per group) dropped out before any follow-up data were collected, leaving 20 analyzed out of 24 randomized. Thus, missing data = 4/24 = 16.7%, which exceeds the 15% threshold. According to the pre-specified thresholds, ≥15% missing data corresponds to “Definitely high.”

Article: Friedman, 2018
 Title: A Randomized, Double-Blind, Placebo-Controlled Trial of Naproxen With or Without Orphenadrine or Methocarbamol for Acute Low Back Pain
 Journal: Annals of Emergency Medicine
 Outcome: Improvement in the Roland-Morris Disability Questionnaire (RMDQ) score between ED discharge and one-week follow-up

### **ITEM 1 (Random sequence generation)**

Step 1
 “Definitely yes”

Step 2
 “Definitely low”

Support for judgment for Item 1
 The trial explicitly states that “The research pharmacist performed randomization in blocks of six based on a sequence generated at [http://randomization.com,”](about:blank) indicating use of an adequate random-number generator for sequence generation .

### **ITEM 2 (Allocation concealment)**

Step 1
 “Definitely yes”

Step 2
 “Definitely low”

Support for judgment for Item 2
 Allocation was centrally managed by the research pharmacist who generated the blocked sequence and masked all investigational tablets by placing them into identical capsules in a secure location inaccessible to ED personnel, ensuring that neither patients nor enrolling clinicians could foresee assignments .

### **ITEM 3 (Blinding of participants)**

Step 1
 “Definitely yes”

Support for judgment for Item 3, Step 1
 The trial is described as “double-blind,” and all orphenadrine, methocarbamol, and placebo tablets were masked in identical capsules; participants therefore could not distinguish active medication from placebo .

Step 2
 “Definitely low”

Support for judgment for Item 3, Step 2
 Because participants were definitively blinded to allocation and the primary outcome (RMDQ) is a self-reported functional measure, blinding would only minimize expectation bias; unblinding is thus very unlikely to have influenced the RMDQ outcome.

### **ITEM 4 (Blinding of healthcare providers)**

Step 1
 “Definitely yes”

Support for judgment for Item 4, Step 1
 The trial is termed “double-blind,” and investigational medications (orphenadrine, methocarbamol, and placebo) were masked in identical capsules by the pharmacist; thus, treating clinicians did not know which muscle relaxant (or placebo) patients received .

Step 2
 “Definitely low”

Support for judgment for Item 4, Step 2
 Healthcare providers were definitively blinded to the specific investigational assignment, and co-interventions were standardized (all patients received naproxen and the same educational session), so unblinding could not have influenced clinical management or the primary functional outcome.

### **ITEM 5 (Blinding of outcome assessors)**

Step 1
 “Definitely yes”

Support for judgment for Item 5, Step 1
 The manuscript states, “The research associates, who were blinded to assignment, collected all of the data using structured interviews” for the one-week RMDQ assessment .

Step 2
 “Definitely low”

Support for judgment for Item 5, Step 2
 Outcome assessors were definitively blinded, and the RMDQ is a standardized questionnaire with minimal subjective interpretation by the assessor, making it very unlikely that unblinding could influence scoring.

### **ITEM 6 (Outcome data not included in analysis)**

Step 1 (Extract the number of participants who were not included in analysis in each group)

From Figure 1 (CONSORT flow diagram) and one-week outcomes:

Intervention group → Naproxen + placebo

- Number of participants randomized: 80
- Number of participants analyzed for one-week RMDQ: 79
- Number not analyzed: 1
- % not analyzed: 1/80 × 100% = 1.25%

Intervention group → Naproxen + orphenadrine

- Number of participants randomized: 80
- Number analyzed: 80
- Number not analyzed: 0
- % not analyzed: 0%

Intervention group → Naproxen + methocarbamol

- Number of participants randomized: 80
- Number analyzed: 81 is reported for one-week outcomes; however, because 240 total were randomized (80 per arm) and Table 2 shows n=81 for methocarbamol, this reflects an inadvertent counting of one extra (likely due to drop-in from other arms). Conservatively, 80 randomized, 81 reported suggests 0 missing, or possibly 0/80 = 0%.
- For consistency with reported denominators (n=81), assume: Number not analyzed: 0; % not analyzed: 0%

Overall →

- Number randomized: 240
- Number analyzed: 79 + 80 + 81 = 240
- Number not analyzed: 0
- % not analyzed: 0%

Step 2
 “Definitely low”

Support for judgment for Item 6, Step 2
 Overall, missing data for the primary outcome (one-week RMDQ) were < 5% in each arm (placebo: 1.25%; orphenadrine: 0%; methocarbamol: 0%), meeting the threshold for “Definitely low” risk (missing < 5%).

Article: Yang, 2024
 Title: COVID-19 Rebound After VV116 vs Nirmatrelvir-Ritonavir Treatment: A Randomized Clinical Trial
 Journal: JAMA Network Open
 Outcome: Viral load rebound (VLR)

### **ITEM 1 (Random sequence generation)**

Step 1
 “Probably yes”

Step 2
 “Probably low”

Support for judgment for Item 1
 The Methods state: “Block randomization with a block size of 4 was used to randomize eligible participants in a 1:1 ratio to receive either VV116 or nirmatrelvir-ritonavir” (). This indicates that participants were described as randomized and that the trial used block randomization, satisfying the “Probably yes” criterion. Because block randomization is an adequate method of sequence generation (even though the exact random number generator is not specified), the risk of bias is “Probably low.”

### **ITEM 2 (Allocation concealment)**

Step 1
 “Probably no”

Step 2
 “Probably high”

Support for judgment for Item 2
 There is no description in the paper of how the allocation sequence was concealed (e.g., central randomization, opaque sealed envelopes). The Methods simply note block randomization without details on concealment (). Because no sufficient information is provided about allocation concealment, this meets the criteria for “Probably no.” In a drug trial without clear concealment, this introduces a “Probably high” risk of bias.

### **ITEM 3 (Blinding of participants)**

Step 1
 “Definitely no”

Support for judgment for Item 3, Step 1
 The Methods specify: “Participants and study staff who were responsible for study drug distribution and recycling were not blinded to the treatment assignment” (). There is no mention of placebo-control or participant masking, so participants were explicitly unblinded.

Step 2
 “Probably low”

Support for judgment for Item 3, Step 2
 Though participants were not blinded, the primary outcome (viral load rebound) is an objective laboratory measurement. Unblinding of participants is unlikely to influence viral load quantification. Hence, risk of bias due to lack of participant blinding is “Probably low.”

### **ITEM 4 (Blinding of healthcare providers)**

Step 1
 “Definitely yes”

Support for judgment for Item 4, Step 1
 The Methods state: “Study investigators, including study physicians who treated participants and collected data … were blinded to the treatment assignment” (). This confirms that healthcare providers responsible for patient care and data collection were blinded.

Step 2
 “Definitely low”

Support for judgment for Item 4, Step 2
 Because healthcare providers were definitely blinded, there is minimal risk that provider unblinding could influence co-interventions or outcome measurement. Therefore, the risk of bias related to provider blinding is “Definitely low.”

### **ITEM 5 (Blinding of outcome assessors)**

Step 1
 “Definitely yes”

Support for judgment for Item 5, Step 1
 The Methods indicate: “Laboratory technicians who tested for viral load … were blinded to the treatment assignment” (). Thus, the personnel performing the primary outcome measurements were explicitly blinded.

Step 2
 “Definitely low”

Support for judgment for Item 5, Step 2
 Measured viral RNA copy number is an objective outcome. Since outcome assessors (laboratory technicians) were definitely blinded and the assay is quantitative, the risk of bias is “Definitely low.”

### **ITEM 6 (Outcome data not included in analysis)**

Step 1 (Extract the number of participants who were not included in analysis in each group)

VV116 group → Number of participants not analyzed: 21
 VV116 group → Total number of participants randomized: 186
 VV116 group → % of participants not analyzed: 11.3%

Nirmatrelvir-ritonavir group → Number of participants not analyzed: 8
 Nirmatrelvir-ritonavir group → Total number of participants randomized: 188
 Nirmatrelvir-ritonavir group → % of participants not analyzed: 4.3%

Overall → Number of participants not analyzed: 29
 Overall → Total number of participants randomized: 374
 Overall → % of participants not analyzed: 7.8%

Step 2
 “Probably low”

Support for judgment for Item 6, Step 2
 Using the pre-specified thresholds (<5% = Definitely low; 5–<10% = Probably low; 10–<15% = Probably high; ≥15% = Definitely high), the overall missing data rate is 7.8%, which falls between 5% and 10%, thus warranting a “Probably low” rating.

Article: Kabon, 2005
 Title: Supplemental Intravenous Crystalloid Administration Does Not Reduce the Risk of Surgical Wound Infection
 Journal: Anesthesia & Analgesia
 Outcome: Incidence of postoperative surgical wound infection

### **ITEM 1 (Random sequence generation)**

Step 1
 “Definitely yes”

Step 2
 “Definitely low”

Support for judgment for Item 1
 The Methods section states that “after induction of anesthesia and endotracheal intubation, patients were assigned to small or large perioperative hydration using computer-generated randomized codes that were kept in opaque, sealed, sequentially numbered envelopes.” This explicitly describes a truly random sequence generated by computer and kept in opaque, sequentially numbered envelopes . Because the trial used an adequate method of random sequence generation (computer-generated codes), risk of bias is “definitely low.”

### **ITEM 2 (Allocation concealment)**

Step 1
 “Definitely yes”

Step 2
 “Definitely low”

Support for judgment for Item 2
 Allocation was concealed by use of “opaque, sealed, sequentially numbered envelopes” containing the computer-generated randomization codes . This approach (sequentially numbered, opaque, sealed envelopes) is explicitly sufficient to prevent foreknowledge of assignment. Therefore, allocation concealment was adequate and risk is “definitely low.”

### **ITEM 3 (Blinding of participants)**

Step 1
 “Definitely no”

Support for judgment for Item 3, Step 1
 The trial is described as open-label with respect to fluid management. There is no mention of participant blinding or use of placebo‐controlled infusion bags; in fact, maintenance rates (8–10 mL·kg⁻¹·h⁻¹ versus 16–18 mL·kg⁻¹·h⁻¹) would have been evident based on the rate of IV infusion . Thus, participants were not blinded.

Step 2
 “Probably high”

Support for judgment for Item 3, Step 2
 Because participants were not blinded (“definitely no”), and knowing infusion rate could influence subjects’ reporting of pain or other postoperative care behaviors, unblinding is likely to have influenced participant‐reported outcomes like pain (VAS) or even perceived wound symptoms. Thus, risk of bias is “probably high.”

### **ITEM 4 (Blinding of healthcare providers)**

Step 1
 “Definitely no”

Support for judgment for Item 4, Step 1
 Anesthesia providers and surgeons necessarily knew infusion rates to manage hemodynamics intraoperatively, and there is no statement that providers were blinded. The clinical team titrated fluids in real time (e.g., “additional fluid was given … to maintain urinary output >1 mL·kg⁻¹·h⁻¹ or when mean arterial pressure decreased …”); thus, providers were aware of group assignment . Consequently, provider blinding was not performed.

Step 2
 “Probably high”

Support for judgment for Item 4, Step 2
 Because healthcare providers were unblinded (“definitely no”), and provider‐initiated co‐interventions (e.g., giving extra fluid boluses, vasopressors, or deciding when to transfuse blood) could influence wound healing and infection risk, unblinding is likely to have influenced the outcome. While the trial standardized many aspects of care (antibiotic use, skin closure technique, wound dressings), fluid administration itself could affect tissue perfusion and oxygenation, and providers may have applied differential co‐interventions (e.g., blood transfusion decisions or hemodynamic adjustments) in each group. This renders risk of bias “probably high.”

### **ITEM 5 (Blinding of outcome assessors)**

Step 1
 “Definitely yes”

Support for judgment for Item 5, Step 1
 The Methods specify that “surgical wounds were evaluated daily by a physician blinded to group assignment. After discharge, the same‐blinded physician evaluated patients … and phone follow‐up was conducted by an investigator blinded to group assignment.” Thus, outcome assessors were explicitly blinded to fluid group .

Step 2
 “Definitely low”

Support for judgment for Item 5, Step 2
 Because outcome assessors were “definitely blinded” and wound infection diagnosis relied on objective CDC criteria (purulent exudate with positive culture or CDC clinical criteria), unblinding could not have influenced assessment. Consequently, risk of bias is “definitely low.”

### **ITEM 6 (Outcome data not included in analysis)**

Step 1 (Extract the number of participants who were not included in analysis in each group)

Intervention group → Number of participants not analyzed: 0
 Intervention group → Total number of participants: 129
 Intervention group → % of participants not analyzed: 0%

Control group → Number of participants not analyzed: 0
 Control group → Total number of participants: 124
 Control group → % of participants not analyzed: 0%

Overall → Number of participants not analyzed: 3
 Overall → Total number of participants: 256
 Overall → % of participants not analyzed: 1.2%

Step 2
 “Definitely low”

Support for judgment for Item 6, Step 2
 Of 256 randomized participants, 253 “completed the trial” . The flow‐chart numbers show that all 124 in the small‐volume group and all 129 in the large‐volume group were analyzed for the main outcome (total n = 253). Thus, there were no missing primary‐outcome data within each arm. Three overall randomized participants withdrew consent before trial completion and were not included in analysis. This 3/256 = 1.2% missing data is well below the 5% threshold for “definitely low.”

**Pre‐specified threshold used:** Missing data < 5% → “definitely low.”

Article: Beretsos, 2009
 Title: “hCG priming” effect in controlled ovarian stimulation through a long protocol
 Journal: Reproductive Biology and Endocrinology
 Outcome: Pregnancy rate

### **ITEM 1 (Random sequence generation)**

Step 1
 Definitely yes

Step 2
 Definitely low

Support for judgment for Item 1
 The Methods section states, “To perform randomization a random number generator<http://www.random.org> was used. For each patient, a random number between 1 and 100,000 was generated and the patient was allocated to the corresponding group (Group 1 for odd numbers and Group 2 for even numbers)” . This clearly describes an adequate, truly random method of sequence generation.

### **ITEM 2 (Allocation concealment)**

Step 1
 Probably no

Step 2
 Probably high

Support for judgment for Item 2
 While the trial details the random sequence generation, there is no description of how allocation was concealed (e.g., no mention of central randomization, sealed envelopes, or pharmacy-controlled assignment). The absence of any statement about concealing the allocation sequence justifies “Probably no.” Because allocation concealment was unclear, this likely introduced a high risk of selection bias (“Probably high”).

### **ITEM 3 (Blinding of participants)**

Step 1
 Definitely no

Support for judgment for Item 3, Step 1
 There is no indication that participants received a placebo or were unaware of their assignment, and pre‐treatment with hCG versus no hCG could not have been masked. The trial does not state that participants were blinded, nor was a “double‐dummy” design used .

Step 2
 Probably low

Support for judgment for Item 3, Step 2
 Although participants were not blinded, the primary outcome—pregnancy rate—is an objective, biologic endpoint (positive serum β-hCG and ultrasound confirmation). Unblinding of participants is unlikely to have influenced this outcome (participant expectations or co-interventions would have minimal impact on implantation and establishment of pregnancy), thus “Probably low.”

### **ITEM 4 (Blinding of healthcare providers)**

Step 1
 Probably no

Support for judgment for Item 4, Step 1
 The study reports that “assessment of follicular growth and endometrial thickness … as well as oocyte collection were performed by the same fertility specialist being unaware of the study group,” and the embryologist evaluating oocyte and embryo quality was also blinded . However, the clinician who initiated hCG pre‐treatment (the treating physician) necessarily knew group assignments. There is no statement that all treating physicians were blinded to allocation, so “Probably no.”

Step 2
 Probably low

Support for judgment for Item 4, Step 2
 Although the clinician administering hCG was aware of group assignment, key co-interventions (e.g., dose adjustments of rFSH) were protocolized and no additional unblinded co-interventions were described. Furthermore, downstream clinical management (ultrasound monitoring, oocyte retrieval) was performed by providers blinded to assignment. Therefore, unblinding of the treating physician is unlikely to have influenced the main outcome (pregnancy rate), justifying “Probably low.”

### **ITEM 5 (Blinding of outcome assessors)**

Step 1
 Definitely yes

Support for judgment for Item 5, Step 1
 The manuscript specifies, “evaluation of oocyte maturation, ICSI procedure and embryo quality assessment were performed by a unique embryologist who was blinded” and “assessment of follicular growth and … oocyte collection … were performed … unaware of the study group in which the patient was assigned” . For pregnancy detection, clinical pregnancy was confirmed by ultrasound two weeks after oocyte retrieval; ultrasound assessors are not described, but pregnancy is an objective measure.

Step 2
 Probably low

Support for judgment for Item 5, Step 2
 Because embryo grading was performed by a blinded embryologist (and pregnancy is determined by objective criteria—serum β-hCG and ultrasound), unblinding of any assessors would be unlikely to influence outcome measurement. Thus, “Probably low.”

### **ITEM 6 (Outcome data not included in analysis)**

Step 1 (Extract the number of participants who were not included in analysis in each group)
 Intervention group → Number of participants not analyzed: 3
 Intervention group → Total number of participants: 22
 Intervention group → % of participants not analyzed: 13.6%

Control group → Number of participants not analyzed: 1
 Control group → Total number of participants: 28
 Control group → % of participants not analyzed: 3.6%

Overall → Number of participants not analyzed: 4
 Overall → Total number of participants: 50
 Overall → % of participants not analyzed: 8.0%

Step 2
 Probably low

Support for judgment for Item 6, Step 2
 The overall missing data (8.0%) falls within the 5%–10% range, which corresponds to “Probably low” according to the pre‐specified thresholds.

Article: Smith, 2017
 Title: A phase I trial of PRN1008, a novel reversible covalent inhibitor of Bruton's tyrosine kinase, in healthy volunteers
 Journal: British Journal of Clinical Pharmacology
 Outcome: Safety, tolerability and pharmacokinetics/pharmacodynamics (BTK occupancy)

### **ITEM 1 (Random sequence generation)**

Step 1
 “Probably yes”

Step 2
 “Probably low”

Support for judgment for Item 1
 The manuscript states, “participants were randomly assigned to PRN1008 or placebo to the next dose cohort” without describing the method used to generate the sequence. This meets the criteria for “Probably yes” (randomized without further details) and “Probably low” (no indication of a quasi‐random method or bias) .

### **ITEM 2 (Allocation concealment)**

Step 1
 “Probably yes”

Step 2
 “Probably low”

Support for judgment for Item 2
 The study is described as “double blind, placebo controlled, randomized” (Part A and Part B) without explicit description of allocation concealment procedures (e.g., opaque envelopes or central allocation). In drug trials where participants and providers are blinded, this is sufficient for “Probably yes” and “Probably low” since no details suggest concealment was inadequate .

### **ITEM 3 (Blinding of participants)**

Step 1
 “Definitely yes”

Support for judgment for Item 3, Step 1
 The Methods state the trial was “double blind, placebo controlled” for both single and multiple ascending dose parts, explicitly indicating that participants were blinded to treatment assignment .

Step 2
 “Definitely low”

Support for judgment for Item 3, Step 2
 Participants’ blinding is certain, and the main outcomes (PK/PD measures like BTK occupancy) are objective laboratory endpoints unlikely to be influenced by participant perception or behavior. Thus, there is no risk of bias from unblinded participants .

### **ITEM 4 (Blinding of healthcare providers)**

Step 1
 “Probably yes”

Support for judgment for Item 4, Step 1
 Although the paper does not explicitly state that healthcare providers were blinded, the description “double blind” implies both participants and providers were unaware of assignment. This qualifies as “Probably yes” in the absence of explicit detail .

Step 2
 “Probably low”

Support for judgment for Item 4, Step 2
 Given that PK/PD and safety endpoints (e.g., laboratory assays, vitals) are objective and not subject to provider‐initiated co‐interventions, even if providers were not definitively blinded, unblinding would be unlikely to influence these outcomes. Therefore, bias risk is “Probably low” .

### **ITEM 5 (Blinding of outcome assessors)**

Step 1
 “Probably yes”

Support for judgment for Item 5, Step 1
 The trial’s “double blind” design suggests that laboratory personnel measuring PK/PD were unaware of treatment assignment, although not explicitly stated. Hence, “Probably yes” applies .

Step 2
 “Probably low”

Support for judgment for Item 5, Step 2
 The primary outcomes—PK parameters (e.g., Cmax, AUC) and BTK occupancy measured via fluorescence assay—are objective laboratory measurements. Even if assessors were not definitively blinded, the objective nature of these assays makes influence unlikely. Therefore, risk is “Probably low” .

### **ITEM 6 (Outcome data not included in analysis)**

Step 1 (Extract the number of participants who were not included in analysis in each group)

Intervention group → Number of participants not analyzed: 0
 Intervention group → Total number of participants: 62
 Intervention group → % of participants not analyzed: 0%

Control group → Number of participants not analyzed: 0
 Control group → Total number of participants: 18
 Control group → % of participants not analyzed: 0%

Overall → Number of participants not analyzed: 0
 Overall → Total number of participants: 80
 Overall → % of participants not analyzed: 0%

Step 2
 “Definitely low”

Support for judgment for Item 6, Step 2
 The paper notes, “All subjects completed the study, and there were no withdrawals or dose interruptions” and “All subjects received their planned doses of study medication” . Missing data are 0%, which is below 5%, thus “Definitely low.”

\# QWEN3-235B-A22B (WITH “THINKING” ACTIVATED) OUTPUT - 31-05-2025

Article: Matsui, 2014

Title: Antibiotic Prophylaxis in Laparoscopic Cholecystectomy: A Randomized Controlled Trial

Journal: PLoS ONE

Outcome: Occurrence of postoperative infections (surgical site infections, distant infections, and overall infections)

\# ITEM 1 (Random sequence generation)

> Step 1

Definitely yes

> Step 2

Definitely low

> Support for judgment for Item 1

The study explicitly stated that randomization was performed using a computer-generated random number just before the operation by a third party (Methods section). This qualifies as an adequate method of random sequence generation (e.g., computer algorithm), fulfilling the criteria for "Definitely yes" and "Definitely low" risk.

\# ITEM 2 (Allocation concealment)

> Step 1

Definitely yes

> Step 2

Definitely low

> Support for judgment for Item 2

Allocation was concealed via central allocation by a third party who telephoned the allocation to the operating theatre (Methods section). Central allocation is explicitly considered a satisfactory concealment method under ROBUST-RCT, warranting "Definitely yes" and "Definitely low" risk.

\# ITEM 3 (Blinding of participants)

> Step 1

Definitely no

> Support for judgment for Item 3, Step 1

The study explicitly stated that the surgeons were not masked to group assignment, and there was no mention of a placebo control. Participants in the No antibiotics group received no intervention, while the Antibiotics group received intravenous cefazolin. These factors indicate open-label design, leading to "Definitely no" for blinding.

> Step 2

Definitely high

> Support for judgment for Item 3, Step 2

Participants were definitely unblinded, and the outcome (postoperative infections) may involve subjective assessments (e.g., wound infections, fever of unknown origin). Participant expectations (e.g., awareness of antibiotic use) could influence reporting of symptoms or adherence to follow-up, leading to "Definitely high" risk.

\# ITEM 4 (Blinding of healthcare providers)

> Step 1

Definitely no

> Support for judgment for Item 4, Step 1

The surgeons providing care were explicitly unblinded to group assignment (Methods section).

> Step 2

Probably high

> Support for judgment for Item 4, Step 2

Healthcare providers were unblinded, and co-interventions (e.g., administration of antibiotics postoperatively for infections) occurred in both groups. While the study mentions standardized protocols for antibiotic administration, the lack of blinding could have influenced decisions regarding additional treatments, leading to "Probably high" risk.

\# ITEM 5 (Blinding of outcome assessors)

> Step 1

Definitely yes

> Support for judgment for Item 5, Step 1

The doctor responsible for outpatient follow-up and infection assessment was unaware of the randomization (Results section).

> Step 2

Definitely low

> Support for judgment for Item 5, Step 2

Outcome assessors were explicitly blinded, and outcomes (e.g., surgical site infections, urinary tract infections) were objectively defined (e.g., pus discharge, microbiological confirmation). This results in "Definitely low" risk.

\# ITEM 6 (Outcome data not included in analysis)

> Step 1

Intervention group → Number of participants not analyzed: 1

Intervention group → Total number of participants: 518

Intervention group → % of participants not analyzed: 0.19%

Control group → Number of participants not analyzed: 2

Control group → Total number of participants: 519

Control group → % of participants not analyzed: 0.39%

Overall → Number of participants not analyzed: 3

Overall → Total number of participants: 1,037

Overall → % of participants not analyzed: 0.29%

> Step 2

Definitely low

> Support for judgment for Item 6, Step 2

Missing data (0.29% overall) is <5%, meeting the "Definitely low" threshold.

---

**Rationale Summary**:

- **Item 1–2**: Adequate randomization and concealment (central allocation).

- **Item 3–4**: No blinding of participants or providers, with potential for bias in subjective outcomes and co-interventions.

- **Item 5**: Outcome assessors were blinded, minimizing detection bias.

- **Item 6**: Minimal missing data (<0.5%), posing negligible bias risk.

Article: Hauser, 2014

Title: Long-term safety and sustained efficacy of extended-release pramipexole in early and advanced Parkinson’s disease

Journal: European Journal of Neurology

Outcome: Unified Parkinson’s Disease Rating Scale (UPDRS) Parts II+III scores

\# ITEM 1 (Random sequence generation)

> Step 1

["Probably no"]

> Step 2

["Probably high"]

> Support for judgment for Item 1

The study describes initial double-blind (DB) trials with randomization but does not explicitly detail the method for generating the random sequence (e.g., random number tables, computer-generated lists). While the DB trials used a double-dummy design, the lack of explicit description of randomization methods (e.g., stratification, block sizes) precludes "Definitely yes." The absence of allocation concealment details further supports "Probably no."

\# ITEM 2 (Allocation concealment)

> Step 1

["Probably yes"]

> Step 2

["Probably low"]

> Support for judgment for Item 2

The DB trials used a double-dummy design (e.g., "IR or placebo as morning, afternoon, and evening doses, plus placebo or ER as a morning dose"). While this implies some allocation concealment, the study does not explicitly confirm sequentially numbered sealed envelopes or central allocation, aligning with "Probably yes" for Step 1. The low risk in Step 2 reflects the assumption that unblinding was unlikely to influence outcomes in the DB phase.

\# ITEM 3 (Blinding of participants)

> Step 1

["Definitely no"]

> Support for judgment for Item 3, Step 1

The study explicitly states that the extension phase was open-label ("OL extension"), meaning participants knew they received pramipexole ER.

> Step 2

["Definitely high"]

> Support for judgment for Item 3, Step 2

The open-label design allowed participant expectations to influence UPDRS Part II (self-reported activities of daily living). The lack of blinding directly impacts the subjective components of the outcome.

\# ITEM 4 (Blinding of healthcare providers)

> Step 1

["Definitely no"]

> Support for judgment for Item 4, Step 1

The OL extension phase did not involve blinding of healthcare providers.

> Step 2

["Definitely high"]

> Support for judgment for Item 4, Step 2

Healthcare providers, aware of the treatment, could influence UPDRS Part III (clinician-rated motor scores) through biased assessments or co-interventions.

\# ITEM 5 (Blinding of outcome assessors)

> Step 1

["Definitely no"]

> Support for judgment for Item 5, Step 1

The study does not mention blinding of outcome assessors in the OL extension phase.

> Step 2

["Definitely high"]

> Support for judgment for Item 5, Step 2

UPDRS Parts II+III involve clinician judgment, particularly Part III (motor examination). Unblinded assessors could introduce bias in scoring, especially for subjective endpoints.

\# ITEM 6 (Outcome data not included in analysis)

> Step 1 (Extract the number of participants who were not included in analysis in each group)

Intervention group → Number of participants not analyzed: 136

Intervention group → Total number of participants: 697

Intervention group → % of participants not analyzed: 19.5%

Control group → Number of participants not analyzed: N/A

Control group → Total number of participants: N/A

Control group → % of participants not analyzed: N/A

Overall → Number of participants not analyzed: 136

Overall → Total number of participants: 697

Overall → % of participants not analyzed: 19.5%

> Step 2

["Definitely high"]

> Support for judgment for Item 6, Step 2

Missing data exceeds 15% (19.5%), triggering the "Definitely high" threshold per pre-specified criteria. Efficacy analyses excluded ex-placebo recipients and had incomplete follow-up (e.g., 234/292 observed cases in early PD OL).

---

**Rationale for missing data calculation**:

- **Early PD (33-week DB)**: 292 ex-DB participants (excluding placebo) had 234 observed cases → 58 missing.

- **Early PD (11–13-week DB)**: 143 OL participants had 113 observed cases → 30 missing.

- **Advanced PD**: 262 ex-DB participants had 214 observed cases → 48 missing.

- **Total missing**: 58 + 30 + 48 = 136.

- **Total analyzed**: 697 participants (435 early PD + 262 advanced PD).

- **Missing proportion**: 136/697 ≈ 19.5%.

Article: Fregni, 2004

Title: Repetitive transcranial magnetic stimulation is as effective as fluoxetine in the treatment of depression in patients with Parkinson’s disease

Journal: Journal of Neurology, Neurosurgery & Psychiatry

Outcome: Hamilton rating scale for depression (HRSD) and Beck depression inventory (BDI) improvement after two weeks of treatment

\# ITEM 1 (Random sequence generation)

> Step 1

Definitely yes

> Step 2

Definitely low

> Support for judgment for Item 1

The study explicitly stated: "The patients were randomly assigned according to a computer generated randomisation list." This qualifies as an adequate method of random sequence generation under ROBUST-RCT criteria.

\# ITEM 2 (Allocation concealment)

> Step 1

Probably yes

> Step 2

Probably low

> Support for judgment for Item 2

The paper did not explicitly describe allocation concealment methods. However, the trial involved a placebo drug (glucose pills) and sham stimulation, with blinding of raters. For drug trials with blinding but unclear concealment details, ROBUST-RCT criteria classify this as "Probably yes/Probably low."

\# ITEM 3 (Blinding of participants)

> Step 1

Definitely yes

> Support for judgment for Item 3, Step 1

The study was described as "double-blind, sham stimulation controlled," and explicitly stated: "a rater blinded to treatment arm." Additionally, blinding integrity was tested by asking interviewers to guess treatment groups.

> Step 2

Definitely low

> Support for judgment for Item 3, Step 2

Participants were definitely blinded (double-blind design). The main outcomes (HRSD, BDI) were clinician-rated, and unblinding was unlikely to influence outcomes due to rater blinding and standardized scales.

\# ITEM 4 (Blinding of healthcare providers)

> Step 1

Probably no

> Support for judgment for Item 4, Step 1

The paper did not explicitly state that healthcare providers (e.g., those administering rTMS or drugs) were blinded. While raters were blinded, providers may have been aware of treatment allocation due to differences in intervention delivery (e.g., active vs. sham rTMS parameters).

> Step 2

Probably low

> Support for judgment for Item 4, Step 2

Although healthcare providers were likely unblinded, the primary outcomes (HRSD, BDI) were assessed by blinded raters. Unblinding was unlikely to influence outcomes because co-interventions were not reported, and standardized assessments were used.

\# ITEM 5 (Blinding of outcome assessors)

> Step 1

Definitely yes

> Support for judgment for Item 5, Step 1

The study explicitly stated: "Psychiatric and neurological evaluation... carried out by raters blinded to the results of the other raters and to the study group assignment."

> Step 2

Definitely low

> Support for judgment for Item 5, Step 2

Outcome assessors were definitely blinded. The main outcomes (HRSD, BDI) involved clinician-rated scales with minimal subjective judgment, and raters were confirmed to be unaware of treatment assignments.

\# ITEM 6 (Outcome data not included in analysis)

> Step 1 (Extract the number of participants who were not included in analysis in each group)

Intervention group → Number of participants not analyzed: 1

Intervention group → Total number of participants: 21

Intervention group → % of participants not analyzed: 4.76%

Control group → Number of participants not analyzed: 0

Control group → Total number of participants: 21

Control group → % of participants not analyzed: 0%

Overall → Number of participants not analyzed: 1

Overall → Total number of participants: 42

Overall → % of participants not analyzed: 2.38%

[If there are more than two arms, adapt the output to extract data for every arm of the study.]

> Step 2

Definitely low

> Support for judgment for Item 6, Step 2

Missing data (2.38%) was <5%, meeting the "Definitely low" threshold per pre-specified criteria. The excluded patient had an unrelated intestinal infection, and all others completed follow-up.

Article: Scarpone, 2008

Title: The efficacy of prolotherapy for lateral epicondylosis: A pilot study

Journal: Clinical Journal of Sport Medicine

Outcome: Resting elbow pain (0–10 Likert scale)

\# ITEM 1 (Random sequence generation)

> Step 1

Definitely yes

> Step 2

Definitely low

> Support for judgment for Item 1

The study explicitly stated: "The 1:1 randomization scheme was prepared by the lead pharmacist... using a random number table and administered using sealed envelopes." This meets the criterion for an adequate random sequence generation method (random number table).

\# ITEM 2 (Allocation concealment)

> Step 1

Definitely yes

> Step 2

Definitely low

> Support for judgment for Item 2

The study used "sealed envelopes" for allocation concealment. While details about sequential numbering or opacity are not fully described, the use of sealed envelopes combined with central randomization by a pharmacist satisfies the criteria for "clearly satisfactory allocation concealment."

\# ITEM 3 (Blinding of participants)

> Step 1

Probably yes

> Support for judgment for Item 3, Step 1

The study was described as "double-blind," and participants received either prolotherapy (active) or saline (placebo). Though not explicitly stated, the use of a placebo-controlled design with identical syringes implies participants were likely blinded.

> Step 2

Probably low

> Support for judgment for Item 3, Step 2

The outcome (resting elbow pain on a 0–10 scale) could be influenced by participant expectations if unblinded. However, since the trial was double-blind and used a placebo control, unblinding was unlikely to have significantly biased the outcome assessment.

\# ITEM 4 (Blinding of healthcare providers)

> Step 1

Definitely yes

> Support for judgment for Item 4, Step 1

The study stated: "Neither prolotherapist nor participant was informed of group status during the study," and syringes were blinded with an opaque sleeve.

> Step 2

Definitely low

> Support for judgment for Item 4, Step 2

Healthcare providers (prolotherapists) were explicitly blinded. There is no evidence of differential co-interventions between groups, further reducing risk of bias.

\# ITEM 5 (Blinding of outcome assessors)

> Step 1

Probably yes

> Support for judgment for Item 5, Step 1

The study was double-blind, and outcome assessors were not explicitly mentioned. However, quantitative measures (e.g., Jamar dynamometer, BTE device) and standardized questionnaires suggest objective assessments.

> Step 2

Definitely low

> Support for judgment for Item 5, Step 2

The primary outcome (resting elbow pain) was self-reported via a Likert scale, making participants the outcome assessors. Since participants were likely blinded, this aligns with "Definitely low" risk. For secondary objective measures (e.g., grip strength), minimal judgment was required.

\# ITEM 6 (Outcome data not included in analysis)

> Step 1

Intervention group → Number of participants not analyzed: 2

Intervention group → Total number of participants: 12

Intervention group → % of participants not analyzed: 16.7%

Control group → Number of participants not analyzed: 2

Control group → Total number of participants: 12

Control group → % of participants not analyzed: 16.7%

Overall → Number of participants not analyzed: 4

Overall → Total number of participants: 24

Overall → % of participants not analyzed: 16.7%

> Step 2

Definitely high

> Support for judgment for Item 6, Step 2

Missing data (16.7%) exceeds the pre-specified threshold of ≥15% ("Definitely high"). Four participants (2 per group) were excluded due to dropout before follow-up data collection.

---

**Key Notes**:

- The study’s primary limitation for Item 6 is the exclusion of 16.7% of randomized participants, which introduces potential selection bias.

- Blinding of participants and providers was rigorously maintained, minimizing performance bias.

- Objective outcome measures reduced detection bias despite partial reliance on self-report.

Article: Friedman, 2018

Title: A Randomized, Double-Blind, Placebo-Controlled Trial of Naproxen With or Without Orphenadrine or Methocarbamol for Acute Low Back Pain

Journal: Annals of Emergency Medicine

Outcome: Improvement on the Roland-Morris Disability Questionnaire (RMDQ) between ED discharge and one week later

\# ITEM 1 (Random sequence generation)

> Step 1

Definitely yes

> Step 2

Definitely low

> Support for judgment for Item 1

The study states: *"The research pharmacist performed randomization in blocks of six based on a sequence generated at http://randomization.com."* This explicit use of a random number generator meets criteria for "Definitely yes" and "Definitely low" risk.

\# ITEM 2 (Allocation concealment)

> Step 1

Definitely yes

> Step 2

Definitely low

> Support for judgment for Item 2

The study describes: *"Orphenadrine, methocarbamol, and placebo were masked by placing tablets into identical capsules [...] packed with [...] sealed. This masking occurred in a secure location inaccessible to ED personnel."* This satisfies "Definitely yes" and "Definitely low" due to clearly described allocation concealment.

\# ITEM 3 (Blinding of participants)

> Step 1

Definitely yes

> Support for judgment for Item 3, Step 1

Participants received identical capsules labeled as "investigational medication," and blinding was confirmed via a follow-up question: *"Do you think you were given the real muscle relaxer or placebo?"* (Appendix 3).

> Step 2

Definitely low

> Support for judgment for Item 3, Step 2

The double-blind design ensured participants were unaware of their allocation. Even though naproxen was unblinded, the primary comparison (add-on effects of orphenadrine/methocarbamol vs. placebo) relied on blinded assessment of subjective outcomes like RMDQ, minimizing bias risk.

\# ITEM 4 (Blinding of healthcare providers)

> Step 1

Definitely yes

> Support for judgment for Item 4, Step 1

ED personnel were excluded from the secure masking process, and study medications were labeled identically: *"bottles [...] labeled as investigational medication."*

> Step 2

Definitely low

> Support for judgment for Item 4, Step 2

Healthcare providers were blinded to treatment allocation, and co-interventions were standardized (e.g., all patients received naproxen and education). No evidence of differential provider-initiated co-interventions was reported.

\# ITEM 5 (Blinding of outcome assessors)

> Step 1

Definitely yes

> Support for judgment for Item 5, Step 1

The study states: *"The research associates, who were blinded to assignment, collected all of the data using structured interviews."*

> Step 2

Definitely low

> Support for judgment for Item 5, Step 2

Outcome assessors were explicitly blinded, and the RMDQ is a standardized self-report questionnaire with yes/no responses. While participant self-report introduces some subjectivity, the structured format reduces interpretation variability.

\# ITEM 6 (Outcome data not included in analysis)

> Step 1

Intervention group → Number of participants not analyzed: 3 (Placebo), 2 (Orphenadrine), 1 (Methocarbamol)

Intervention group → Total number of participants: 79 (Placebo), 80 (Orphenadrine), 81 (Methocarbamol)

Intervention group → % of participants not analyzed: 3.8% (Placebo), 2.5% (Orphenadrine), 1.2% (Methocarbamol)

Control group → N/A (three-arm trial)

Overall → Number of participants not analyzed: 6

Overall → Total number of participants: 240

Overall → % of participants not analyzed: 2.5%

> Step 2

Definitely low

> Support for judgment for Item 6, Step 2

Missing data (2.5%) falls below the 5% threshold for "Definitely low" risk.

Article: Yang, 2024

Title: COVID-19 Rebound After VV116 vs Nirmatrelvir-Ritonavir Treatment: A Randomized Clinical Trial

Journal: JAMA Network Open

Outcome: Viral load rebound (VLR), defined as a half-log increase in viral RNA copies/mL post-treatment completion.

\# ITEM 1 (Random sequence generation)

> Step 1

**Definitely yes**

> Step 2

**Definitely low**

> Support for judgment for Item 1

The trial explicitly stated: "Block randomization with a block size of 4 was used to randomize eligible participants in a 1:1 ratio." This meets the criteria for "Definitely yes" (explicit use of block randomization) and "Definitely low" risk (adequate method).

\# ITEM 2 (Allocation concealment)

> Step 1

**Definitely yes**

> Step 2

**Definitely low**

> Support for judgment for Item 2

The trial used block randomization with a predefined block size and 1:1 allocation ratio. This satisfies "Definitely yes" for explicit allocation concealment and "Definitely low" risk (block randomization ensures concealed allocation).

\# ITEM 3 (Blinding of participants)

> Step 1

**Definitely no**

> Support for judgment for Item 3, Step 1

The paper states: "Participants and study staff who were responsible for study drug distribution and recycling were not blinded to the treatment assignment." Participants were explicitly unblinded.

> Step 2

**Probably low**

> Support for judgment for Item 3, Step 2

The primary outcome (VLR) is objective (viral load measured via PCR/ddPCR). Participant unblinding is unlikely to influence this outcome, though the non-blinded design could introduce bias in subjective measures (not applicable here).

\# ITEM 4 (Blinding of healthcare providers)

> Step 1

**Definitely no**

> Support for judgment for Item 4, Step 1

The study explicitly states that "participants and study staff responsible for drug distribution were not blinded."

> Step 2

**Probably low**

> Support for judgment for Item 4, Step 2

The primary outcome (VLR) is objective and not influenced by healthcare provider actions. No co-interventions were documented to differ between groups.

\# ITEM 5 (Blinding of outcome assessors)

> Step 1

**Definitely yes**

> Support for judgment for Item 5, Step 1

The paper states: "laboratory technicians who tested for viral load... were blinded to the treatment assignment."

> Step 2

**Definitely low**

> Support for judgment for Item 5, Step 2

Outcome assessors (lab technicians) were blinded, and the primary outcome (VLR) was measured via standardized PCR/ddPCR, which involves minimal subjective judgment.

\# ITEM 6 (Outcome data not included in analysis)

> Step 1

Intervention group → Number of participants not analyzed: **21** (186 enrolled, 165 analyzed)

Intervention group → Total number of participants: **186**

Intervention group → % of participants not analyzed: **11.3%**

Control group → Number of participants not analyzed: **8** (188 enrolled, 180 analyzed)

Control group → Total number of participants: **188**

Control group → % of participants not analyzed: **4.3%**

Overall → Number of participants not analyzed: **29** (374 randomized, 345 analyzed)

Overall → Total number of participants: **374**

Overall → % of participants not analyzed: **7.8%**

> Step 2

**Probably low**

> Support for judgment for Item 6, Step 2

Missing data (7.8%) falls within the pre-specified threshold of 5% ≤ missing data <10% ("Probably low"). Censoring due to missed follow-up was applied uniformly for time-to-event analyses.

Article: Kabon, 2005

Title: Supplemental Intravenous Crystalloid Administration Does Not Reduce the Risk of Surgical Wound Infection

Journal: Anesthesia & Analgesia

Outcome: Surgical wound infection

\# ITEM 1 (Random sequence generation)

> Step 1

Definitely yes

> Step 2

Definitely low

> Support for judgment for Item 1

The study explicitly states: "patients were assigned to small or large perioperative hydration using computer-generated randomized codes that were kept in opaque, sealed, sequentially numbered envelopes." This confirms adequate randomization (e.g., computer-generated).

\# ITEM 2 (Allocation concealment)

> Step 1

Definitely yes

> Step 2

Definitely low

> Support for judgment for Item 2

The allocation sequence was concealed using "opaque, sealed, sequentially numbered envelopes," and the anesthesia records were sealed until 16 days post-surgery, ensuring blinding of surgeons and outcome assessors.

\# ITEM 3 (Blinding of participants)

> Step 1

Probably no

> Support for judgment for Item 3, Step 1

The study does not explicitly state that participants were blinded. The intervention (fluid volume) was administered intravenously, making participant blinding unlikely without explicit mention.

> Step 2

Probably low

> Support for judgment for Item 3, Step 2

The primary outcome (surgical wound infection) is objective and less susceptible to bias from participant expectations.

\# ITEM 4 (Blinding of healthcare providers)

> Step 1

Definitely no

> Support for judgment for Item 4, Step 1

The anesthesiologists managing fluid administration were aware of group assignments, as stated: "anesthesia records and perioperative fluid administration records were sealed... Thus, the surgeons and investigators evaluating wound infections... were unable to determine group assignment." Providers were not explicitly blinded.

> Step 2

Probably low

> Support for judgment for Item 4, Step 2

Additional fluid administration was guided by predefined criteria (urinary output, blood pressure), minimizing provider discretion. Co-interventions (e.g., antibiotics) were standardized, reducing bias risk.

\# ITEM 5 (Blinding of outcome assessors)

> Step 1

Definitely yes

> Support for judgment for Item 5, Step 1

"The surgical wounds were evaluated daily by a physician blinded to group assignment. After discharge, the same-blinded physician evaluated patients... patients not returning to the clinic were contacted by phone by an investigator blinded to group assignment."

> Step 2

Definitely low

> Support for judgment for Item 5, Step 2

Outcome assessors were explicitly blinded, and the primary outcome (wound infection) was assessed using objective criteria (e.g., purulent exudate + culture, CDC criteria).

\# ITEM 6 (Outcome data not included in analysis)

> Step 1

Intervention group → Number of participants not analyzed: 0

Intervention group → Total number of participants: 129

Intervention group → % of participants not analyzed: 0%

Control group → Number of participants not analyzed: 3

Control group → Total number of participants: 124

Control group → % of participants not analyzed: 2.4%

Overall → Number of participants not analyzed: 3

Overall → Total number of participants: 256

Overall → % of participants not analyzed: 1.2%

> Step 2

Definitely low

> Support for judgment for Item 6, Step 2

Missing data (1.2%) is <5%, meeting the "Definitely low" threshold.

---

**Final Summary:**

- **Item 1:** Definitely low risk

- **Item 2:** Definitely low risk

- **Item 3:** Probably low risk

- **Item 4:** Probably low risk

- **Item 5:** Definitely low risk

- **Item 6:** Definitely low risk

The study demonstrates robust methodology with minimal risk of bias across all ROBUST-RCT domains.

Article: Beretsos, 2009

Title: "hCG priming effect in controlled ovarian stimulation through a long protocol"

Journal: Reproductive Biology and Endocrinology

Outcome: Pregnancy rate

\# ITEM 1 (Random sequence generation)

> Step 1

["Definitely yes"]

> Step 2

["Definitely low"]

> Support for judgment for Item 1

The study explicitly states: "To perform randomization a random number generator [http://www.random.org] was used. For each patient, a random number between 1 and 100,000 was generated and the patient was allocated to the corresponding group (Group 1 for odd numbers and Group 2 for even numbers)." This confirms the use of a valid randomization method (random number generator), fulfilling "Definitely yes" for Step 1 and "Definitely low" risk for Step 2.

\# ITEM 2 (Allocation concealment)

> Step 1

["Probably no"]

> Step 2

["Probably high"]

> Support for judgment for Item 2

The paper does not describe any allocation concealment method (e.g., sealed envelopes, central allocation). While randomization was performed via random.org, there is no mention of how the group assignments were concealed from investigators during enrollment. This falls under "Probably no" for Step 1 and "Probably high" risk for Step 2.

\# ITEM 3 (Blinding of participants)

> Step 1

["Definitely no"]

> Support for judgment for Item 3, Step 1

The study does not explicitly state that participants were blinded. Given the intervention (hCG pre-treatment) and control protocols differ in treatment timing and drug administration, participants would likely be aware of their group assignment.

> Step 2

["Probably low"]

> Support for judgment for Item 3, Step 2

Pregnancy rate (main outcome) is objectively confirmed by ultrasound (gestational sac with fetal heartbeat), minimizing the influence of participant expectations on outcome assessment. Thus, unblinding is unlikely to significantly bias results.

\# ITEM 4 (Blinding of healthcare providers)

> Step 1

["Definitely yes"]

> Support for judgment for Item 4, Step 1

The Methods section states: "Assessment of follicular growth and endometrial thickness by ultrasound scan... were performed by the same fertility specialist being unaware of the study group... Similarly, evaluation of oocyte maturation, ICSI procedure, and embryo quality assessment were performed by a unique embryologist who was blinded too."

> Step 2

["Definitely low"]

> Support for judgment for Item 4, Step 2

Healthcare providers (fertility specialist and embryologist) were explicitly blinded, reducing risk of bias in procedural execution and outcome assessment.

\# ITEM 5 (Blinding of outcome assessors)

> Step 1

["Definitely yes"]

> Support for judgment for Item 5, Step 1

As noted in Item 4, the fertility specialist and embryologist were blinded to group assignments during outcome assessments (e.g., follicular growth, embryo quality, pregnancy confirmation via ultrasound).

> Step 2

["Definitely low"]

> Support for judgment for Item 5, Step 2

Pregnancy confirmation via ultrasound (gestational sac with fetal heartbeat) is an objective outcome assessed by blinded personnel, fulfilling "Definitely low" risk.

\# ITEM 6 (Outcome data not included in analysis)

> Step 1

Intervention group → Number of participants not analyzed: [3]

Intervention group → Total number of participants: [22]

Intervention group → % of participants not analyzed: [13.6]%

Control group → Number of participants not analyzed: [1]

Control group → Total number of participants: [28]

Control group → % of participants not analyzed: [3.6]%

Overall → Number of participants not analyzed: [4]

Overall → Total number of participants: [50]

Overall → % of participants not analyzed: [8]%

> Step 2

["Probably low"]

> Support for judgment for Item 6, Step 2

Missing data (8%) falls within the 5–10% threshold ("Probably low" risk), as pre-specified in the guidance.

Article: Smith, 2017

Title: A phase I trial of PRN1008, a novel reversible covalent inhibitor of Bruton’s tyrosine kinase, in healthy volunteers

Journal: British Journal of Clinical Pharmacology

Outcome: Safety (adverse events) and pharmacodynamic effect (BTK occupancy)

\# ITEM 1 (Random sequence generation)

> Step 1

Probably yes

> Step 2

Probably low

> Support for judgment for Item 1

The study described as "randomized" but did not specify the method of random sequence generation (e.g., random number tables, computer-generated sequences). It mentioned "double-blind, placebo-controlled, randomized" and provided allocation ratios (e.g., "6 active/2 placebo per cohort"), but no details on randomization procedures. This meets "Probably yes" for Step 1 and "Probably low" for Step 2 under quasi-randomization criteria.

\# ITEM 2 (Allocation concealment)

> Step 1

Probably no

> Step 2

Probably high

> Support for judgment for Item 2

The paper did not explicitly describe allocation concealment methods (e.g., sealed envelopes, central allocation). Participants were assigned to "next dose cohort" after eligibility confirmation, but no details on how allocations were concealed. As this is a drug trial with unclear concealment, this aligns with "Probably no" for Step 1 and "Probably high" for Step 2.

\# ITEM 3 (Blinding of participants)

> Step 1

Definitely yes

> Support for judgment for Item 3, Step 1

The study was explicitly described as "double-blind" (Methods section: "double blind, placebo-controlled, randomized"). For a placebo-controlled drug trial, this satisfies "Definitely yes" for participant blinding.

> Step 2

Definitely low

> Support for judgment for Item 3, Step 2

Participants were definitely blinded ("Definitely yes" for Step 1), and the primary outcome (safety/adverse events) is unlikely to be influenced by participant expectations. Objective measures (e.g., laboratory parameters) were prioritized, meeting "Definitely low" risk.

\# ITEM 4 (Blinding of healthcare providers)

> Step 1

Definitely yes

> Support for judgment for Item 4, Step 1

The study was described as "double-blind," implying blinding of both participants and healthcare providers. No evidence contradicts this assumption.

> Step 2

Definitely low

> Support for judgment for Item 4, Step 2

Healthcare providers were definitely blinded ("Definitely yes" for Step 1). There were no reported provider-initiated co-interventions, and safety assessments relied on standardized protocols (e.g., adverse event monitoring, laboratory tests), minimizing risk of bias.

\# ITEM 5 (Blinding of outcome assessors)

> Step 1

Probably no

> Support for judgment for Item 5, Step 1

While the study was "double-blind," the paper did not explicitly state whether outcome assessors (e.g., those evaluating adverse events or BTK occupancy) were blinded. For non-drug trials or unclear reporting, this aligns with "Probably no."

> Step 2

Probably high

> Support for judgment for Item 5, Step 2

Outcome assessors were not explicitly confirmed as blinded ("Probably no" for Step 1). Adverse events (a primary outcome) involve subjective reporting, and BTK occupancy measurements, though objective, required adjudication. Unblinding could plausibly influence outcome assessment, leading to "Probably high" risk.

\# ITEM 6 (Outcome data not included in analysis)

> Step 1

Intervention group → Number of participants not analyzed: 0

Intervention group → Total number of participants: 62

Intervention group → % of participants not analyzed: 0%

Control group → Number of participants not analyzed: 0

Control group → Total number of participants: 18

Control group → % of participants not analyzed: 0%

Overall → Number of participants not analyzed: 0

Overall → Total number of participants: 80

Overall → % of participants not analyzed: 0%

> Step 2

Definitely low

> Support for judgment for Item 6, Step 2

All 80 participants completed the study with no withdrawals or missing data ("All subjects completed the study, and there were no withdrawals or dose interruptions"). Missing data = 0%, satisfying the "Definitely low" threshold.
