## Appendix 3 for "Assessment of Bias in Clinical Trials with LLMs Using ROBUST-RCT: A Feasibility Study"

```
COEFFICIENTS


GPT-4-turbo

dimension    method    coefficient    stderr    p_value
item1step1    Gwet_AC2    0.86033    0.12542    0.0001297287304
item1step2    Gwet_AC2    0.86033    0.12542    0.0001297287304
item2step1    Gwet_AC2    0.51497    0.33985    0.168162862
item2step2    Gwet_AC2    0.51497    0.33985    0.168162862
item3step1    Gwet_AC2    0.51171    0.29082    0.1165224064
item3step2    Gwet_AC2    0.42531    0.32639    0.2287993914
item4step1    Gwet_AC2    0.31075    0.32991    0.3738033851
item4step2    Gwet_AC2    0.43509    0.21524    0.07788591542
item5step1    Gwet_AC2    0.8125     0.16195    0.001030767486
item5step2    Gwet_AC2    0.66165    0.26104    0.03499717444
item6step2    Gwet_AC2    0.77186    0.17009    0.001904500295


Gemini 2.5 Pro Preview

dimension    method    coefficient    stderr    p_value
item1step1    Gwet_AC2    0.90155    0.0637     6.04E-07
item1step2    Gwet_AC2    0.90155    0.0637     6.04E-07
item2step1    Gwet_AC2    0.58916    0.33044    0.1124416747
item2step2    Gwet_AC2    0.58916    0.33044    0.1124416747
item3step1    Gwet_AC2    0.78082    0.09239    2.94E-05
item3step2    Gwet_AC2    0.55       0.1946     0.02227546613
item4step1    Gwet_AC2    0.53064    0.29802    0.1128515439
item4step2    Gwet_AC2    0.52381    0.20822    0.03605197786
item5step1    Gwet_AC2    0.81772    0.12804    0.0002121238825
item5step2    Gwet_AC2    0.77028    0.13681    0.0004924805461
item6step2    Gwet_AC2    0.81226    0.16414    0.001122726099


DeepThink R1

dimension    method    coefficient    stderr    p_value
item1step1    Gwet_AC2    0.90155    0.05211    1.27E-07
item1step2    Gwet_AC2    0.90155    0.05211    1.27E-07
item2step1    Gwet_AC2    0.48645    0.33617    0.185907204
item2step2    Gwet_AC2    0.48645    0.33617    0.185907204
item3step1    Gwet_AC2    0.2063     0.32949    0.5486799277
item3step2    Gwet_AC2    0.25311    0.16879    0.1721071345
item4step1    Gwet_AC2   -0.02395    0.44824    0.9586949521
item4step2    Gwet_AC2    0.3527     0.14718    0.04342671427
item5step1    Gwet_AC2    0.52673    0.31483    0.1328470942
item5step2    Gwet_AC2    0.33657    0.37182    0.3918065584
item6step2    Gwet_AC2    0.77186    0.17009    0.001904500295


Qwen3-235B-A22B

dimension    method    coefficient    stderr    p_value
item1step1    Gwet_AC2    0.90155    0.0637     6.04E-07
item1step2    Gwet_AC2    0.90155    0.0637     6.04E-07
item2step1    Gwet_AC2    0.46846    0.33257    0.1966126259
item2step2    Gwet_AC2    0.46846    0.33257    0.1966126259
item3step1    Gwet_AC2    0.48645    0.29821    0.1414846238
item3step2    Gwet_AC2    0.66316    0.26148    0.034920893
item4step1    Gwet_AC2    0.03933    0.39247    0.9226501629
item4step2    Gwet_AC2    0.60049    0.15264    0.004332687702
item5step1    Gwet_AC2    0.55828    0.30565    0.1051942381
item5step2    Gwet_AC2    0.63236    0.2763     0.05136673088
item6step2    Gwet_AC2    0.77186    0.17009    0.001904500295
```

```
BENCHMARKING


GPT-4-turbo

item1step1
                Landis-Koch CumProb
(0.8 to 1)   Almost Perfect 0.63651
(0.6 to 0.8)    Substantial 0.97814
(0.4 to 0.6)       Moderate 0.99986
(0.2 to 0.4)           Fair       1
(0 to 0.2)           Slight       1
(-1 to 0)              Poor       1

item1step2
                Landis-Koch CumProb
(0.8 to 1)   Almost Perfect 0.63651
(0.6 to 0.8)    Substantial 0.97814
(0.4 to 0.6)       Moderate 0.99986
(0.2 to 0.4)           Fair       1
(0 to 0.2)           Slight       1
(-1 to 0)              Poor       1

item2step1
                Landis-Koch CumProb
(0.8 to 1)   Almost Perfect 0.13437
(0.6 to 0.8)    Substantial 0.35143
(0.4 to 0.6)       Moderate 0.60187
(0.2 to 0.4)           Fair 0.80827
(0 to 0.2)           Slight 0.92976
(-1 to 0)              Poor       1


item2step2
                Landis-Koch CumProb
(0.8 to 1)   Almost Perfect 0.13437
(0.6 to 0.8)    Substantial 0.35143
(0.4 to 0.6)       Moderate 0.60187
(0.2 to 0.4)           Fair 0.80827
(0 to 0.2)           Slight 0.92976
(-1 to 0)              Poor       1


item3step1
                Landis-Koch CumProb
(0.8 to 1)   Almost Perfect 0.11977
(0.6 to 0.8)    Substantial 0.35047
(0.4 to 0.6)       Moderate 0.63244
(0.2 to 0.4)           Fair 0.85117
(0 to 0.2)           Slight 0.95884
(-1 to 0)              Poor       1


item3step2
                Landis-Koch CumProb
(0.8 to 1)   Almost Perfect 0.08987
(0.6 to 0.8)    Substantial 0.26758
(0.4 to 0.6)       Moderate  0.5118
(0.2 to 0.4)           Fair 0.74502
(0 to 0.2)           Slight 0.89981
(-1 to 0)              Poor       1


item4step1
                Landis-Koch CumProb
(0.8 to 1)   Almost Perfect 0.05164
(0.6 to 0.8)    Substantial 0.17519
(0.4 to 0.6)       Moderate 0.38205
(0.2 to 0.4)           Fair 0.62459
(0 to 0.2)           Slight 0.82368
(-1 to 0)              Poor       1


item4step2
                Landis-Koch CumProb
(0.8 to 1)   Almost Perfect 0.04084
(0.6 to 0.8)    Substantial  0.2184
(0.4 to 0.6)       Moderate 0.56286
(0.2 to 0.4)           Fair 0.86203
(0 to 0.2)           Slight 0.97829
(-1 to 0)              Poor       1


item5step1
                Landis-Koch CumProb
(0.8 to 1)   Almost Perfect 0.46466
(0.6 to 0.8)    Substantial 0.89192
(0.4 to 0.6)       Moderate  0.9938
(0.2 to 0.4)           Fair 0.99991
(0 to 0.2)           Slight       1
(-1 to 0)              Poor       1


item5step2
                Landis-Koch CumProb
(0.8 to 1)   Almost Perfect 0.22226
(0.6 to 0.8)    Substantial 0.54944
(0.4 to 0.6)       Moderate 0.82484
(0.2 to 0.4)           Fair 0.95736
(0 to 0.2)           Slight 0.99376
(-1 to 0)              Poor       1


item6step2
               Landis-Koch CumProb
(0.8 to 1)   Almost Perfect 0.37841
(0.6 to 0.8)    Substantial 0.82842
(0.4 to 0.6)       Moderate 0.98418
(0.2 to 0.4)           Fair 0.99958
(0 to 0.2)           Slight       1
(-1 to 0)              Poor       1


Gemini 2.5 Pro Preview

item1step1
                Landis-Koch CumProb
(0.8 to 1)   Almost Perfect 0.94094
(0.6 to 0.8)    Substantial       1
(0.4 to 0.6)       Moderate       1
(0.2 to 0.4)           Fair       1
(0 to 0.2)           Slight       1
(-1 to 0)              Poor       1


item1step2
                Landis-Koch CumProb
(0.8 to 1)   Almost Perfect 0.94094
(0.6 to 0.8)    Substantial       1
(0.4 to 0.6)       Moderate       1
(0.2 to 0.4)           Fair       1
(0 to 0.2)           Slight       1
(-1 to 0)              Poor       1


item2step1
                Landis-Koch CumProb
(0.8 to 1)   Almost Perfect 0.17337
(0.6 to 0.8)    Substantial 0.42552
(0.4 to 0.6)       Moderate 0.68257
(0.2 to 0.4)           Fair 0.86625
(0 to 0.2)           Slight 0.95824
(-1 to 0)              Poor       1


item2step2
                Landis-Koch CumProb
(0.8 to 1)   Almost Perfect 0.17337
(0.6 to 0.8)    Substantial 0.42552
(0.4 to 0.6)       Moderate 0.68257
(0.2 to 0.4)           Fair 0.86625
(0 to 0.2)           Slight 0.95824
(-1 to 0)              Poor       1

item3step1
                Landis-Koch CumProb
(0.8 to 1)   Almost Perfect 0.41258
(0.6 to 0.8)    Substantial 0.97461
(0.4 to 0.6)       Moderate 0.99998
(0.2 to 0.4)           Fair       1
(0 to 0.2)           Slight       1
(-1 to 0)              Poor       1


item3step2
                Landis-Koch CumProb
(0.8 to 1)   Almost Perfect 0.09001
(0.6 to 0.8)    Substantial 0.39231
(0.4 to 0.6)       Moderate 0.77728
(0.2 to 0.4)           Fair 0.96358
(0 to 0.2)           Slight 0.99762
(-1 to 0)              Poor       1

item4step1
                Landis-Koch CumProb
(0.8 to 1)   Almost Perfect 0.13308
(0.6 to 0.8)    Substantial 0.37177
(0.4 to 0.6)       Moderate 0.64922
(0.2 to 0.4)           Fair 0.85821
(0 to 0.2)           Slight 0.96021
(-1 to 0)              Poor       1


item4step2
                Landis-Koch CumProb
(0.8 to 1)   Almost Perfect 0.08216
(0.6 to 0.8)    Substantial    0.35
(0.4 to 0.6)       Moderate 0.72085
(0.2 to 0.4)           Fair 0.93937
(0 to 0.2)           Slight 0.99399
(-1 to 0)              Poor       1


item5step1
                Landis-Koch CumProb
(0.8 to 1)   Almost Perfect 0.51777
(0.6 to 0.8)    Substantial 0.95174
(0.4 to 0.6)       Moderate  0.9994
(0.2 to 0.4)           Fair       1
(0 to 0.2)           Slight       1
(-1 to 0)              Poor       1


item5step2
                Landis-Koch CumProb
(0.8 to 1)   Almost Perfect 0.38539
(0.6 to 0.8)    Substantial 0.88816
(0.4 to 0.6)       Moderate 0.99643
(0.2 to 0.4)           Fair 0.99998
(0 to 0.2)           Slight       1
(-1 to 0)              Poor       1


item6step2
                Landis-Koch CumProb
(0.8 to 1)   Almost Perfect 0.46176
(0.6 to 0.8)    Substantial 0.88785
(0.4 to 0.6)       Moderate 0.99312
(0.2 to 0.4)           Fair 0.99989
(0 to 0.2)           Slight       1
(-1 to 0)              Poor       1


DeepThink R1

item1step1
                Landis-Koch CumProb
(0.8 to 1)   Almost Perfect 0.97356
(0.6 to 0.8)    Substantial       1
(0.4 to 0.6)       Moderate       1
(0.2 to 0.4)           Fair       1
(0 to 0.2)           Slight       1
(-1 to 0)              Poor       1

item1step2
                Landis-Koch CumProb
(0.8 to 1)   Almost Perfect 0.97356
(0.6 to 0.8)    Substantial       1
(0.4 to 0.6)       Moderate       1
(0.2 to 0.4)           Fair       1
(0 to 0.2)           Slight       1
(-1 to 0)              Poor       1

item2step1

                Landis-Koch CumProb
(0.8 to 1)   Almost Perfect 0.11977
(0.6 to 0.8)    Substantial 0.32504
(0.4 to 0.6)       Moderate 0.57454
(0.2 to 0.4)           Fair 0.78961
(0 to 0.2)           Slight 0.92106
(-1 to 0)              Poor       1


item2step2
                Landis-Koch CumProb
(0.8 to 1)   Almost Perfect 0.11977
(0.6 to 0.8)    Substantial 0.32504
(0.4 to 0.6)       Moderate 0.57454
(0.2 to 0.4)           Fair 0.78961
(0 to 0.2)           Slight 0.92106
(-1 to 0)              Poor       1


item3step1
                Landis-Koch CumProb
(0.8 to 1)   Almost Perfect 0.02801
(0.6 to 0.8)    Substantial 0.10895
(0.4 to 0.6)       Moderate 0.27252
(0.2 to 0.4)           Fair 0.50372
(0 to 0.2)           Slight 0.73233
(-1 to 0)              Poor       1

item3step2
                Landis-Koch CumProb
(0.8 to 1)   Almost Perfect 0.00059
(0.6 to 0.8)    Substantial 0.01993
(0.4 to 0.6)       Moderate 0.19208
(0.2 to 0.4)           Fair 0.62348
(0 to 0.2)           Slight 0.93313
(-1 to 0)              Poor       1

item4step1
                Landis-Koch CumProb
(0.8 to 1)   Almost Perfect 0.02242
(0.6 to 0.8)    Substantial 0.07267
(0.4 to 0.6)       Moderate 0.16523
(0.2 to 0.4)           Fair 0.30541
(0 to 0.2)           Slight 0.47995
(-1 to 0)              Poor       1


item4step2
                Landis-Koch CumProb
(0.8 to 1)   Almost Perfect 0.00118
(0.6 to 0.8)    Substantial 0.04645
(0.4 to 0.6)       Moderate 0.37396
(0.2 to 0.4)           Fair 0.85025
(0 to 0.2)           Slight 0.99172
(-1 to 0)              Poor       1

item5step1
                Landis-Koch CumProb
(0.8 to 1)   Almost Perfect 0.13529
(0.6 to 0.8)    Substantial 0.36589
(0.4 to 0.6)       Moderate 0.63192
(0.2 to 0.4)           Fair 0.83968
(0 to 0.2)           Slight 0.94949
(-1 to 0)              Poor       1

item5step2
                Landis-Koch CumProb
(0.8 to 1)   Almost Perfect  0.0718
(0.6 to 0.8)    Substantial 0.20997
(0.4 to 0.6)       Moderate 0.41041
(0.2 to 0.4)           Fair 0.62963
(0 to 0.2)           Slight  0.8104
(-1 to 0)              Poor       1

item6step2
                Landis-Koch CumProb
(0.8 to 1)   Almost Perfect 0.37841
(0.6 to 0.8)    Substantial 0.82842
(0.4 to 0.6)       Moderate 0.98418
(0.2 to 0.4)           Fair 0.99958
(0 to 0.2)           Slight       1
(-1 to 0)              Poor       1


Qwen3-235B-A22B

item1step1
                Landis-Koch CumProb
(0.8 to 1)   Almost Perfect 0.94094
(0.6 to 0.8)    Substantial       1
(0.4 to 0.6)       Moderate       1
(0.2 to 0.4)           Fair       1
(0 to 0.2)           Slight       1
(-1 to 0)              Poor       1

item1step2
                Landis-Koch CumProb
(0.8 to 1)   Almost Perfect 0.94094
(0.6 to 0.8)    Substantial       1
(0.4 to 0.6)       Moderate       1
(0.2 to 0.4)           Fair       1
(0 to 0.2)           Slight       1
(-1 to 0)              Poor       1

item2step1
                Landis-Koch CumProb
(0.8 to 1)   Almost Perfect 0.11049
(0.6 to 0.8)    Substantial 0.30819
(0.4 to 0.6)       Moderate  0.5572
(0.2 to 0.4)           Fair 0.77803
(0 to 0.2)           Slight  0.9159
(-1 to 0)              Poor       1

item2step2
                Landis-Koch CumProb
(0.8 to 1)   Almost Perfect 0.11049
(0.6 to 0.8)    Substantial 0.30819
(0.4 to 0.6)       Moderate  0.5572
(0.2 to 0.4)           Fair 0.77803
(0 to 0.2)           Slight  0.9159
(-1 to 0)              Poor       1

item3step1
                Landis-Koch CumProb
(0.8 to 1)   Almost Perfect 0.10862
(0.6 to 0.8)    Substantial 0.32289
(0.4 to 0.6)       Moderate 0.59691
(0.2 to 0.4)           Fair 0.82414
(0 to 0.2)           Slight  0.9463
(-1 to 0)              Poor       1

item3step2
                Landis-Koch CumProb
(0.8 to 1)   Almost Perfect 0.22364
(0.6 to 0.8)    Substantial 0.55106
(0.4 to 0.6)       Moderate 0.82566
(0.2 to 0.4)           Fair 0.95755
(0 to 0.2)           Slight 0.99378
(-1 to 0)              Poor       1

item4step1
                Landis-Koch CumProb
(0.8 to 1)   Almost Perfect 0.01933
(0.6 to 0.8)    Substantial 0.07016
(0.4 to 0.6)       Moderate 0.17382
(0.2 to 0.4)           Fair 0.33774
(0 to 0.2)           Slight 0.53878
(-1 to 0)              Poor       1

item4step2
                Landis-Koch CumProb
(0.8 to 1)   Almost Perfect 0.09157
(0.6 to 0.8)    Substantial 0.49906
(0.4 to 0.6)       Moderate 0.90507
(0.2 to 0.4)           Fair 0.99563
(0 to 0.2)           Slight 0.99996
(-1 to 0)              Poor       1

item5step1
                Landis-Koch CumProb
(0.8 to 1)   Almost Perfect 0.15156
(0.6 to 0.8)    Substantial 0.40129
(0.4 to 0.6)       Moderate 0.67349
(0.2 to 0.4)           Fair 0.86978
(0 to 0.2)           Slight  0.9634
(-1 to 0)              Poor       1

item5step2
                Landis-Koch CumProb
(0.8 to 1)   Almost Perfect 0.19855
(0.6 to 0.8)    Substantial 0.50086
(0.4 to 0.6)       Moderate 0.77962
(0.2 to 0.4)           Fair 0.93525
(0 to 0.2)           Slight 0.98784
(-1 to 0)              Poor       1

item6step2
                Landis-Koch CumProb
(0.8 to 1)   Almost Perfect 0.37841
(0.6 to 0.8)    Substantial 0.82842
(0.4 to 0.6)       Moderate 0.98418
(0.2 to 0.4)           Fair 0.99958
(0 to 0.2)           Slight       1
(-1 to 0)              Poor       1
```
