## Supplementary Table 1 for "Assessment of Bias in Clinical Trials with LLMs Using ROBUST-RCT: A Feasibility Study"

**Supplementary Table 1.** Pre-specified exclusion criteria.

| **Reason** | **Description** |
| --- | --- |
| 1. Not an RCT | Studies that are not randomized controlled trials (e.g., single-arm studies, systematic reviews, case reports, poster presentations, randomized controlled trial protocol without results). |
| 2. Not individually randomized | Studies that are not individually randomized human trials (e.g., cluster-randomized trials, crossover designs, animal studies). |
| 3. Not the original study | Post-hoc analyses and secondary analyses that were not the original primary purpose and main outcome of the study design, even if planned before the trial. |
| 4. Other | Other studies for which ROBUST-RCT analysis is not feasible or useful (e.g., lack of access to the full article, language isn’t English). |
