## Supplementary Table 2 for "Assessment of Bias in Clinical Trials with LLMs Using ROBUST-RCT: A Feasibility Study"

**Supplementary Table 2.** Possible answers for the ROBUST-RCT assessment. Step 1 (except item 6) – DY: definitely yes; PY: probably yes; PN: probably no; DN: definitely no. Step 2 – DL: definitely low; PL: probably low; PH: probably high; DH: definitely high.

| Item | Step 1 | Step 2 |
| --- | --- | --- |
| 1 – Random sequence generation | Was the allocation sequence adequately generated?  **DY / PY / PN / DN** | Judge risk of bias related to sequence generation  **DL / PL / PH / DH** |
| 2 – Allocation concealment | Was the allocation adequately concealed?  **DY / PY / PN / DN** | Judge risk of bias related to allocation concealment  **DL / PL / PH / DH** |
| 3 – Blinding of participants | Were participants blinded?  **DY / PY / PN / DN** | Judge risk of bias related to blinding of participants  **DL / PL / PH / DH** |
| 4 – Blinding of healthcare providers | Were healthcare providers blinded?  **DY / PY / PN / DN** | Judge risk of bias related to blinding of healthcare providers  **DL / PL / PH / DH** |
| 5 – Blinding of outcome assessors | Were outcome assessors blinded?  **DY / PY / PN / DN** | Judge risk of bias related to blinding of outcome assessors  **DL / PL / PH / DH** |
| 6 – Outcome data not included in analysis | Extract the number of participants who were not included in analysis in each group. *(Open-ended; not evaluated directly in our data analysis)* | Judge risk of bias related to the overall percentage of participants not included in analysis.  **DL / PL / PH / DH** |
