## Supplementary Table 3 for "Assessment of Bias in Clinical Trials with LLMs Using ROBUST-RCT: A Feasibility Study"

**Supplementary Table 3.** Pre-specified thresholds for item 6 of ROBUST-RCT.

| Definitely low | Missing data < 5% |
| --- | --- |
| Probably low | 5% ≤ Missing data < 10% |
| Probably high | 10% ≤ Missing data < 15% |
| Definitely high | Missing data ≥ 15% |
