## Supplementary Table 4 for "Assessment of Bias in Clinical Trials with LLMs Using ROBUST-RCT: A Feasibility Study"

**Supplementary Table 4.** RCTs included in the pilot.

| **Title** | **First Author** | **Year** | **Journal** | **PMCID** |
| --- | --- | --- | --- | --- |
| Antibiotic prophylaxis in laparoscopic cholecystectomy: a randomized controlled trial | Matsui Y | 2014 | PLoS One | PMC4156368 |
| Long-term safety and sustained efficacy of extended-release pramipexole in early and advanced Parkinson's disease | Hauser RA | 2014 | Eur J Neurol | PMC4282380 |
| Repetitive transcranial magnetic stimulation is as effective as fluoxetine in the treatment of depression in patients with Parkinson's disease | Fregni F | 2004 | J Neurol Neurosurg Psychiatry | PMC1739189 |
| The efficacy of prolotherapy for lateral epicondylosis: a pilot study | Scarpone M | 2008 | Clin J Sport Med | PMC2751593 |
| A Randomized, Double-Blind, Placebo-Controlled Trial of Naproxen With or Without Orphenadrine or Methocarbamol for Acute Low Back Pain | Friedman BW | 2018 | Ann Emerg Med | PMC5820149 |
| COVID-19 Rebound After VV116 vs Nirmatrelvir-Ritonavir Treatment: A Randomized Clinical Trial | Yang Z | 2024 | JAMA Netw Open | PMC10938176 |
| Supplemental intravenous crystalloid administration does not reduce the risk of surgical wound infection | Kabon B | 2005 | Anesth Analg | PMC1388094 |
| hCG priming effect in controlled ovarian stimulation through a long protocol | Beretsos P | 2009 | Reprod Biol Endocrinol | PMC2744681 |
| A phase I trial of PRN1008, a novel reversible covalent inhibitor of Bruton's tyrosine kinase, in healthy volunteers | Smith PF | 2017 | Br J Clin Pharmacol | PMC5651318 |
