## Supplementary Table 5 for "Assessment of Bias in Clinical Trials with LLMs Using ROBUST-RCT: A Feasibility Study"

**Supplementary Table 5.** Fleiss’ Kappa among the three reviewers (before consensus).

| **Fleiss’ Kappa** | **Percentage of Agreement** | **Percentage Expected** | **Standard Error** | **95% Confidence Interval** |
| --- | --- | --- | --- | --- |
| 0.4941 | 0.6666 | 0.3410 | 0.0947 | 0.304, 0.684 |
