## Supplementary Table 6 for "Assessment of Bias in Clinical Trials with LLMs Using ROBUST-RCT: A Feasibility Study"

**Supplementary Table 7.** Mean ordinal values for each group. Higher values indicate a stricter assessment (risk of bias towards "definitely high"), and lower values suggest a more lenient assessment (risk of bias towards "definitely low").

| **Reviewer** | **Mean** | **Standard Deviation** |
| --- | --- | --- |
| Human consensus | 0.808 | 0.997 |
| GPT-4-turbo | 0.990 | 1.15 |
| Gemini 2.5 Pro Preview | 0.758 | 0.905 |
| DeepSeek-R1 | 1.16 | 1.07 |
| Qwen3-235B-A22B | 0.859 | 1.12 |
