## Supplementary Table 7 for "Assessment of Bias in Clinical Trials with LLMs Using ROBUST-RCT: A Feasibility Study"

**Supplementary Table 8.** Direction of bias analysis. Wilcoxon Signed-Rank test. DeepSeek had the two sides of the confidence interval with negative values, which suggests it's a more stringent reviewer in comparison to the human consensus.

| **Comparison** | **95% Confidence Interval (lower)** | **95% Confidence Interval (upper)** | **p-value** | **adjusted p-value (Bonferroni)** |
| --- | --- | --- | --- | --- |
| GPT-4-turbo | -1.000048046236446053214e+00 | 5.452279760143264502117e-05 | 0.1085 | 0.4343 |
| Gemini 2.5 Pro Preview | -6.548702838840233814849e-06 | 9.999754820407968924911e-01 | 0.6467 | 1 |
| DeepSeek-R1 | -1.000038431374988601164e+00 | -1.087794978974344732581e-06 | **0.0050*** | **0.0201*** |
| Qwen3-235B-A22B | -0.9999569516477651021091 | 0.4999819470143812560892 | 0.7159 | 1 |
| ***Significance** | | | | |
